# GWAS and ExWAS of blood Mitochondrial DNA copy number identifies 73 loci and highlights a potential causal role in dementia

**DOI:** 10.1101/2021.04.08.21255031

**Authors:** Michael Chong, Pedrum Mohammadi-Shemirani, Nicolas Perrot, Walter Nelson, Robert W. Morton, Sukrit Narula, Ricky Lali, Irfan Khan, Mohammad Khan, Conor Judge, Tafadzwa Machipisa, Nathan Cawte, Martin O’Donnell, Marie Pigeyre, Loubna Akhabir, Guillaume Paré

## Abstract

Mitochondrial DNA copy number (mtDNA-CN) is an accessible blood-based measurement believed to capture underlying mitochondrial function. The specific biological processes underpinning its regulation, and whether those processes are causative for disease, is an area of active investigation. We developed a novel method for array-based mtDNA-CN estimation suitable for biobank-scale studies, called “AutoMitoC”. We applied AutoMitoC to 395,781 UKBiobank study participants and performed genome and exome-wide association studies, identifying novel common and rare genetic determinants. Overall, genetic analyses identified 73 loci for mtDNA-CN, which implicated several genes involved in rare mtDNA depletion disorders, dNTP metabolism, and the mitochondrial central dogma. Rare variant analysis identified *SAMHD1* mutation carriers as having higher mtDNA-CN (beta=0.23 SDs; 95% CI, 0.18-0.29; P=2.6×10^−19^), a potential therapeutic target for patients with mtDNA depletion disorders, but at increased risk of breast cancer (OR=1.91; 95% CI, 1.52-2.40; P=2.7×10^−8^). Finally, Mendelian randomization analyses suggest a causal effect of low mtDNA-CN on dementia risk (OR=1.94 per 1 SD decrease in mtDNA-CN; 95% CI, 1.55-2.32; P=7.5×10^−4^). Altogether, our genetic findings indicate that mtDNA-CN is a complex biomarker reflecting specific mitochondrial processes related to mtDNA regulation, and that these processes are causally related to human diseases.

## Introduction

Mitochondria are semi-autonomous organelles present in nearly every human cell that execute fundamental cellular processes including oxidative phosphorylation, calcium storage, and apoptotic signalling. Mitochondrial dysfunction has been implicated as the underlying cause for many human disorders based on mechanistic *in vitro* and *in vivo* studies (Burbulla et al. 2017; Desdín-micó et al. 2020; Sliter et al. 2020). Complementary evidence comes from recent epidemiological studies that measure mitochondrial DNA Copy Number (mtDNA-CN), a marker of mitochondrial activity that can be conveniently measured from peripheral blood. Since mitochondria contain their own unique set of genomes that are distinct from the nuclear genome, the ratio of mtDNA to nuclear DNA molecules (mtDNA-CN) in a sample serves as an accessible marker of mitochondrial quantity (Ryan J. Longchamps et al. 2020). Indeed, observational studies suggest that individuals with lower mtDNA-CN are at higher risk of age-related complex diseases, such as coronary artery disease, sudden cardiac death, cardiomegaly, stroke, portal hypertension, and chronic kidney disease (Ashar et al. 2017; Hägg et al. 2020; Tin et al. 2016; Zhang et al. 2017). Conversely, higher mtDNA-CN levels have been associated with increased cancer incidence (Hu, Yao, and Shen 2016; Kim et al. 2015).

While previous studies demonstrate that mtDNA-CN is a biomarker of mitochondrial activity associated with various diseases, evidence suggests that it may also play a direct and causative role in human health and disease. For example, in cases of mtDNA depletion syndrome, wherein rare defects in nuclear genes responsible for replicating and/or maintaining mtDNA lead to deficient mtDNA-CN (Gorman et al. 2016), patients manifest with severe dysfunction of energy-dependent tissues (heart, brain, liver, and cardiac and skeletal muscles). So far, 19 genes have been reported to cause mtDNA depletion (Oyston 1998). In addition to these rare monogenic syndromes, the importance of common genetic variation in regulating mtDNA-CN is an active area of research with approximately 50 common loci identified so far (Cai et al. 2015; Guyatt et al. 2019; Hägg et al. 2020; Ryan Joseph Longchamps 2019).

To interrogate mtDNA-CN as a potential determinant of human diseases, we performed extensive genetic investigations in up to 395,781 participants from the UKBiobank study(Sudlow et al. 2015). We first developed and validated a novel method for biobank-scale mtDNA-CN investigations that leverages SNP array intensities, called “AutoMitoC”. Leveraging AutoMitoC-based mtDNA-CN estimates, we performed large-scale GWAS and ExWAS to identify common and rare genetic variants contributing to population-level variation in mtDNA-CN. Various analyses were then conducted to build on previous publications regarding the specific genes and pathways underlying mtDNA-CN regulation(Cai et al. 2015; Guyatt et al. 2019; Hägg et al. 2020; Ryan Joseph Longchamps 2019). Finally, we applied Mendelian randomization analyses to assess potential causal relationships between mtDNA-CN and disease susceptibility.

## Methods

### The UKBiobank study

The UKBiobank is a prospective cohort study including approximately 500,000 UK residents (ages 40-69 years) recruited from 2006-2010 in whom extensive genetic and phenotypic investigations have been and continue to be done(Sudlow et al. 2015). All UKBiobank data reported in this manuscript were accessed through the UKBiobank data showcase under application #15525. All analyses involve the use of genetic and/or phenotypic data from consenting UKBiobank participants.

### Genetic Analysis of Common Variants

#### Data acquisition and quality control

Imputed genotypes (version 3) for 488,264 UKBiobank participants were downloaded through the European Genome Archive (Category 100319). Detailed sample and variant quality control are described in the supplementary methods. In special consideration of mtDNA-CN as the GWAS phenotype, we also removed variants within “NUMTs”, which refer to regions of the nuclear genome that exhibit homology to the mitochondrial genome due to past transposition of mitochondrial sequences (Simone et al. 2011). After quality control, 359,689 British, 10,598 Irish, 13,189 Other White, 6,172 South Asian, and 6,133 African samples had suitable array-based mtDNA-CN estimates for subsequent GWAS testing.

#### Association testing

GWAS were initially conducted in an ethnicity-stratified manner for common variants (MAF > 0.005). To allow for genetic relatedness between participants, GWAS were conducted using the REGENIE framework (Mbatchou et al. 2020). GWAS covariates included age, age^2^, sex, chip type, 20 genetic principal components, and blood cell counts (white blood cell, platelet, and neutrophil counts). After ethnicity-specific GWAS were performed, results were combined through meta-analysis using METAL (Willer, Li, and Abecasis 2010). European (N=383,476) and trans-ethnic (N=395,718) GWAS meta-analyses were performed. See supplementary methods for further details.

#### Fine-mapping of GWAS signals

We followed a similar protocol to Vuckovic *et al*. (2020) for fine-mapping mtDNA-CN loci (Vuckovic et al. 2020). Genome-wide significant variants were consolidated into genomic blocks by grouping variants within 250kb of each other. LDstore was used to compute a pairwise LD correlation matrix for all variants within each block and across all samples included in the European GWAS meta-analysis (Benner et al. 2017). For each genomic block, FINEMAP was used to perform stepwise conditional regression (Benner et al. 2016). The number of conditionally independent genetic signals per genomic block was used to inform the subsequent fine-mapping search parameters. Finally, the FINEMAP random stochastic search algorithm was applied to derive 95% credible sets constituting candidate causal variants that jointly contributed to 95% (or higher) of the posterior inclusion probabilities (Benner et al. 2016).

#### Mitochondrial expression quantitative trait loci (mt-eQTL) and heteroplasmy look-ups

Among GWAS hits, we searched for mt-eQTLs using information from Ali *et al*. (2019), “Nuclear genetic regulation of the human mitochondrial transcriptome”(Ali et al. 2019). All variants in both Tables 1 and 2 were queried in the mtDNA-CN summary statistics. When mt-eQTLs also had reported effect estimates, the consistency in direction-of-effects between mt-eQTL and mtDNA-CN associations was reported (S2. Table 3). GWS variants from a heteroplasmy GWAS by Nandakumar *et al*. (2021)(Nandakumar et al. 2021).

#### Gene prioritization & pathway analyses

The Data-driven Expression-prioritized Integration for Complex Traits (DEPICT) v.1.1 tool was used to map mtDNA-CN loci to genes based on shared co-regulation of gene expression using default settings (Pers et al. 2015). DEPICT-prioritized genes were uploaded to the GeneMANIA web platform (https://genemania.org/). Based on the combined list of DEPICT and GeneMANIA identified genes, a network was formed in GeneMANIA using default settings. Functional enrichment analysis was then performed to identify overrepresented Gene Ontology (GO) terms among all network genes (Gene and Consortium 2000).

#### Mitochondrial annotation-based analyses

To complement the previous pathway analyses, we labelled prioritized genes with MitoCarta3 annotations and performed subsequent statistical enrichment analyses (Rath et al. 2021). MitoCarta3 is an exquisite database of mitochondrial protein annotations, which draws from mass spectrophotometry and GFP colocalization experiments of isolated mitochondria from 14 different tissues to assign all human genes statuses indicating whether the corresponding proteins are expressed in the mitochondria or not. We tested whether prioritized genes were enriched for the mitochondrial proteome by using a binomial test in R. Furthermore, a t-test was used to compare mean PGC-1A induced fold change for the subset of GWAS-prioritized genes expressed in the mitochondrial proteome as compared to the mean PGC-1A induced fold change for all 1120 nuclear MitoCarta3-annotated genes. Also, genes were categorized based on MitoCarta3 “MitoPathway” annotations.

### Genetic Analysis of Rare Variants

#### Data acquisition and quality control

Population-level whole-exome sequencing (WES) variant genotypes (UKB data field: 23155) for 200,643 UKBiobank participants corresponding to 17,975,236 variants were downloaded using the gfetch utility. Detailed quality control of WES data is described in the supplementary methods. After quality control, 173,688 unrelated Caucasian samples remained.

#### Exome-wide association study (ExWAS) to identify rare mtDNA-CN loci

Of the 173,688 individuals, suitable AutoMitoC mtDNA-CN estimates were available for 147,740 samples. Rare variant inclusion criteria consisted of variants which were infrequent (MAF ≤ 0.001), non-synonymous, and predicted to be clinically deleterious by Mendelian Clinically Applicable Pathogenicity (M-CAP) v.1.4 scores (or were highly disruptive variant types including frameshift indel, stopgain, stoploss, or splicing) (Jagadeesh et al. 2016). Herein, such variants are referred to as “rare variants” for simplicity. For each gene, rare allele counts were added per sample. A minor allele count of 10 was applied leading to a total of 18,890 genes analyzed (exome-wide significance P < 0.05/18,890 = 2.65 ×10^−6^). Linear regression was conducted using mtDNA-CN as the dependent variable and the rare alleles counts per gene as the independent variable. The same set of covariates used in the primary GWAS analysis was used in the ExWAS analysis.

#### Phenome-wide association testing for rare SAMHD1 mutation carrier status

To identify disease phenotypes associated with carrying a rare *SAMHD1* mutation, we maximized sample size for phenome-wide association testing by analyzing the larger set of 173,688 WES samples (with or without suitable mtDNA-CN estimates). Disease outcomes were defined using the previously published “PheCode” classification scheme to aggregate ICD-10 codes from hospital episodes (field ID 41270), death registry (field ID 40001 and 40002), and cancer registry (field ID 40006) records(Denny et al. 2013; P. Wu et al. 2019). Logistic regression was applied to test the association of *SAMHD1* mutation carrier status versus 771 PheCodes (phenome-wide significance P < 0.05/771 = 6.49×10^−5^) with a minimal case sample size of 300 (Wei et al. 2017). The same set of covariates used in the primary GWAS were also employed in this analysis.

### Mendelian Randomization Analysis

#### Disease Outcomes

We cross-referenced a list of 36 clinical manifestations of mitochondrial disease to FinnGen consortium GWAS traits (Gorman et al. 2016; Madewell et al. 2020). Among 2,444 FinnGen traits, 10 overlapped with mitochondrial disease and had a case prevalence greater than 1% and were chosen for two-sample Mendelian Randomization analyses. These 10 traits included type 2 diabetes (N=23,364), mood disorder (N=20,288), sensorineural hearing loss (N=12,550), cerebrovascular disease (N=10,367), migraine (N=6,687), dementia (5,675), epilepsy (N=4,558), paralytic ileus and intestinal obstruction (N=2,999), and cardiomyopathy (N=2,342). FinnGen effect estimates and standard errors were used in subsequent Mendelian randomization analyses to define the effect of selected genetic instruments on disease risk.

#### Genetic Instrument Selection

First, genome-wide significant variants from the present European GWAS meta-analysis of mtDNA-CN were chosen (N=383,476). Second, we matched these variants to the FinnGen v4 GWAS datasets (Madewell et al. 2020). Third, to enrich for variants that directly act through mitochondrial processes, we only retained those within 100kb of genes encoding for proteins that are expressed in mitochondria based on MitoCarta3 annotations (Rath et al. 2021). Fourth, we performed LD-pruning in PLINK with 1000Genomes Europeans as the reference panel to ascertain an independent set of genetic variants (LD r2 > 0.01) (Abecasis et al. 2012; Purcell et al. 2007). Lastly, to mitigate potential for horizontal pleiotropy, we further removed variants with strong evidence of acting through alternative pathways by performing a phenome-wide search across published GWAS with Phenoscanner V2 (Kamat et al. 2019). Variants strongly associated with other phenotypes (P<5×10^−20^) were removed unless the variant was a coding mutation located within gene encoding for the mitochondrial proteome (MitoCarta3) or had an established mitochondrial role based on manual literature review (Rath et al. 2021). A total of 27 genetic variants were used to approximate genetically determined mtDNA-CN levels.

#### Mendelian Randomization & Sensitivity Analyses

Two sample Mendelian Randomization analyses were performed using the “TwoSampleMR” and “MRPRESSO” R packages (Hemani et al. 2018; Verbanck et al. 2018). Effect estimates and standard errors corresponding to the 27 genetic variants on mtDNA-CN (exposure) and mitochondrial disease phenotypes (outcome) were derived from the European GWAS meta-analysis and FinnGen v4 GWAS summary statistics, respectively (S2. Table 9). Three MR methodologies were employed including Inverse Variance Weighted (primary method), Weighted Median, and MR-EGGER methods. MR-PRESSO was used to detect global heterogeneity and P-values were derived based on 1000 simulations. If significant global heterogeneity was detected (P<0.05), a local outlier test was conducted to detect outlying SNPs. After removal of outlying SNPs, MR analyses were repeated. In the absence of heterogeneity (Egger-intercept P ≥ 0.05; MR-PRESSO global heterogeneity P ≥ 0.05), we reported the inverse-variance weighted result. In the presence of balanced pleiotropy (MR-PRESSO global heterogeneity P < 0.05) and absence of directional pleiotropy (Egger-intercept P ≥ 0.05), we reported the weighted median result. In the presence of directional pleiotropy (Egger-intercept P < 0.05), we reported the MR-EGGER result.

## Results

### AutoMitoC: A streamlined method for array-based mtDNA-CN estimation

We built on an existing framework for processing normalized SNP probe intensities (L2R values) from genetic arrays into mtDNA-CN estimates known as the “Mitopipeline” (Lane 2014). The MitoPipeline yields mtDNA-CN estimates that correlate with direct qPCR measurements and has been successfully implemented in several epidemiological investigations(Ashar et al. 2017; Zhang et al. 2017). We developed a novel method, “AutoMitoC”, which incorporates three amendments to facilitate large-scale investigations of mtDNA-CN. Firstly, AutoMitoC replaces autosomal signal normalization of common variants with globally rare variants which negates the need for linkage disequilibrium pruning. As a result, this simplifies derivation of mtDNA-CN estimates in ethnically diverse cohorts by allowing for use of a single, universal variant set for normalization. Secondly, to detect potentially cross-hybridizing probes, we empirically assess the association of corrected probe signal intensities with off-target genome intensities, rather than relying on sequence homology of probe sequences, which is not always available. Lastly, the primary estimate of MT signal is ascertained using principal component analysis (as opposed to using the median signal intensity of MT probes as per the Mitopipeline) which improves concordance of array-based mtDNA-CN estimates with those derived from alternative methods. A detailed description of the AutoMitoC pipeline is provided in Supplementary Results 1.

To benchmark performance of AutoMitoC, array-based mtDNA-CN estimates were compared to complementary measures of mtDNA-CN in two independent studies. Firstly, array-based mtDNA-CN estimates were derived in a subset of 34,436 UKBiobank participants with available whole exome sequencing (WES) data. Reference mtDNA-CN estimates were derived from the proportion of WES reads aligned to the mitochondrial genome relative to the autosome (Ryan Joseph Longchamps 2019). AutoMitoC estimates were significantly correlated with WES estimates (r=0.45; P<2.23×10^−308^). Since WES data involves enrichment for nuclear coding genes and therefore could result in biased reference estimates for mtDNA-CN, we also performed an independent validation in an ethnically diverse study of 5,791 participants where mtDNA-CN was measured using quantitative PCR, the current gold standard assay (Fazzini et al. 2018). Indeed, we observed stronger correlation between AutoMitoC and qPCR-based estimates (r=0.64; P<2.23×10^−308^). Furthermore, AutoMitoC demonstrated robust performance (r ≥ 0.53) across all ethnic strata in the secondary validation cohort including Europeans (N=2,431), Latin Americans (N=1,704), Africans (N=542), South East Asians (N=471), South Asians (N=186), and others (N=360; S1. Figure 4).

### Genome-wide association study (GWAS) identifies 72 common loci for mtDNA-CN

A GWAS was performed testing the association between 11,453,766 common genetic variants (MAF>0.005) with mtDNA-CN in 383,476 UKBiobank participants of European ancestry. In total, 9,602 variants were associated with mtDNA-CN at genome-wide significance (Figure 1A; S2. Figure 1), encompassing 82 independent signals in 72 loci (S2. Table 1; S2. Figure 2). The genomic inflation factor was 1.16 and the LD-score intercept was 1.036, indicating that most inflation in test statistics was attributable to polygenicity.

**Figure 1.**
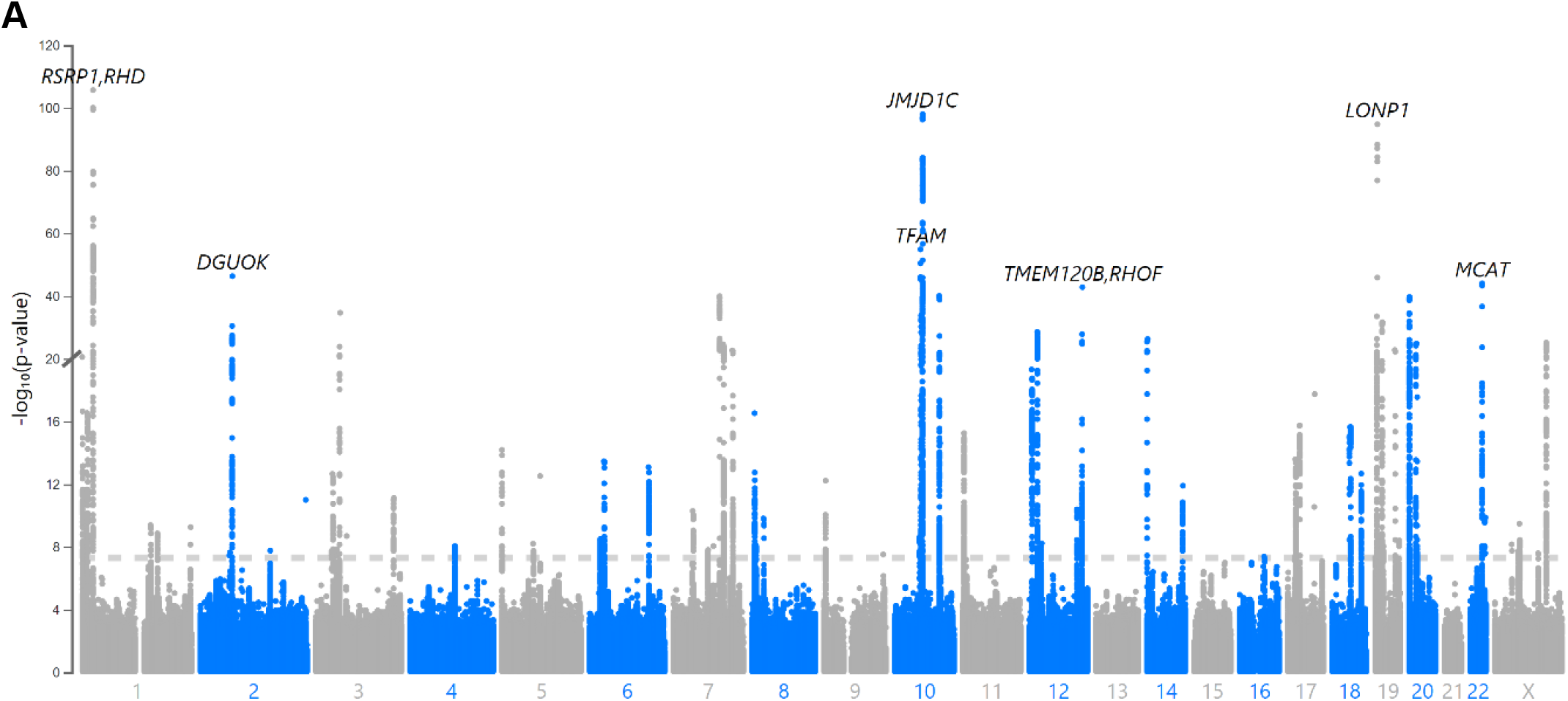
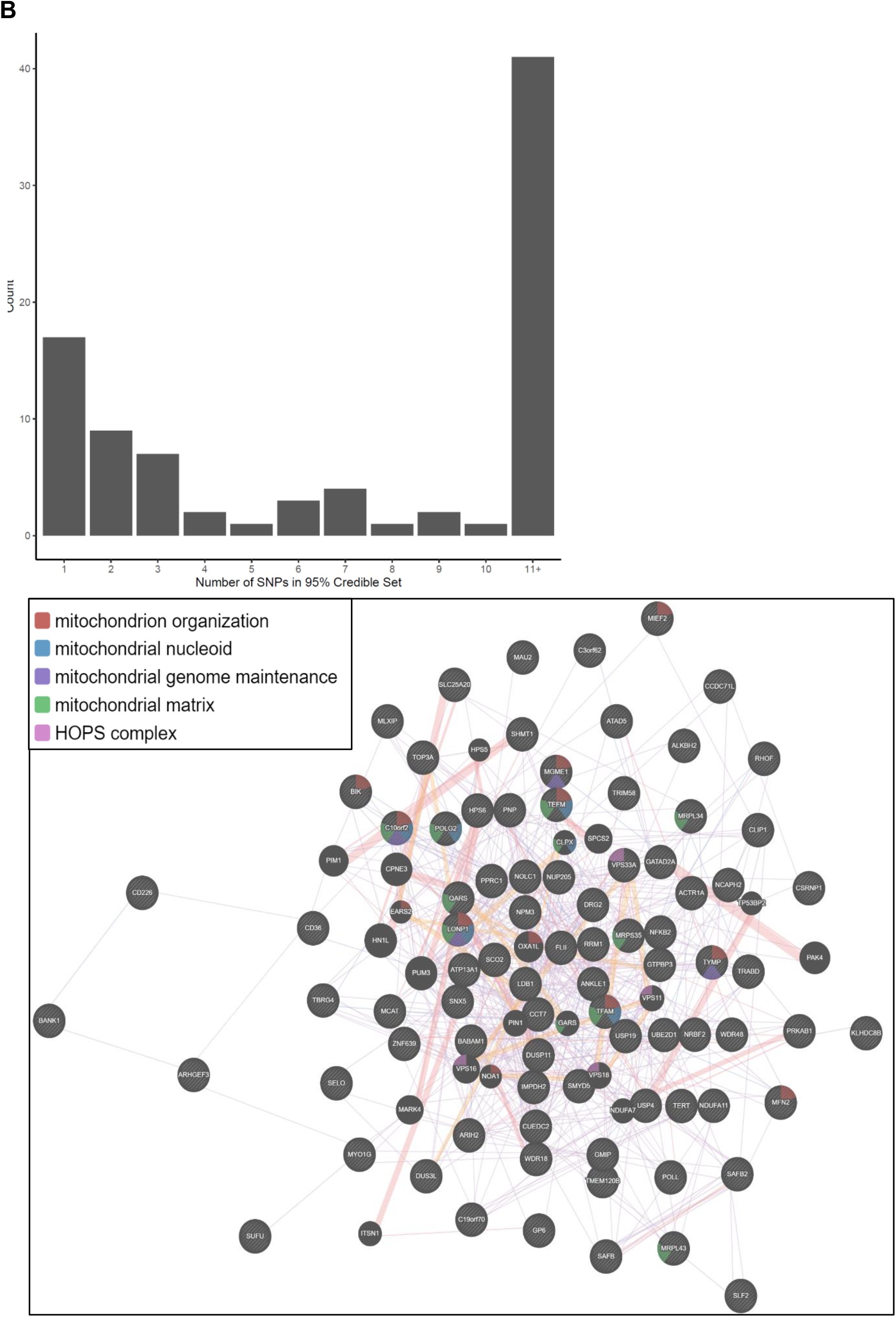
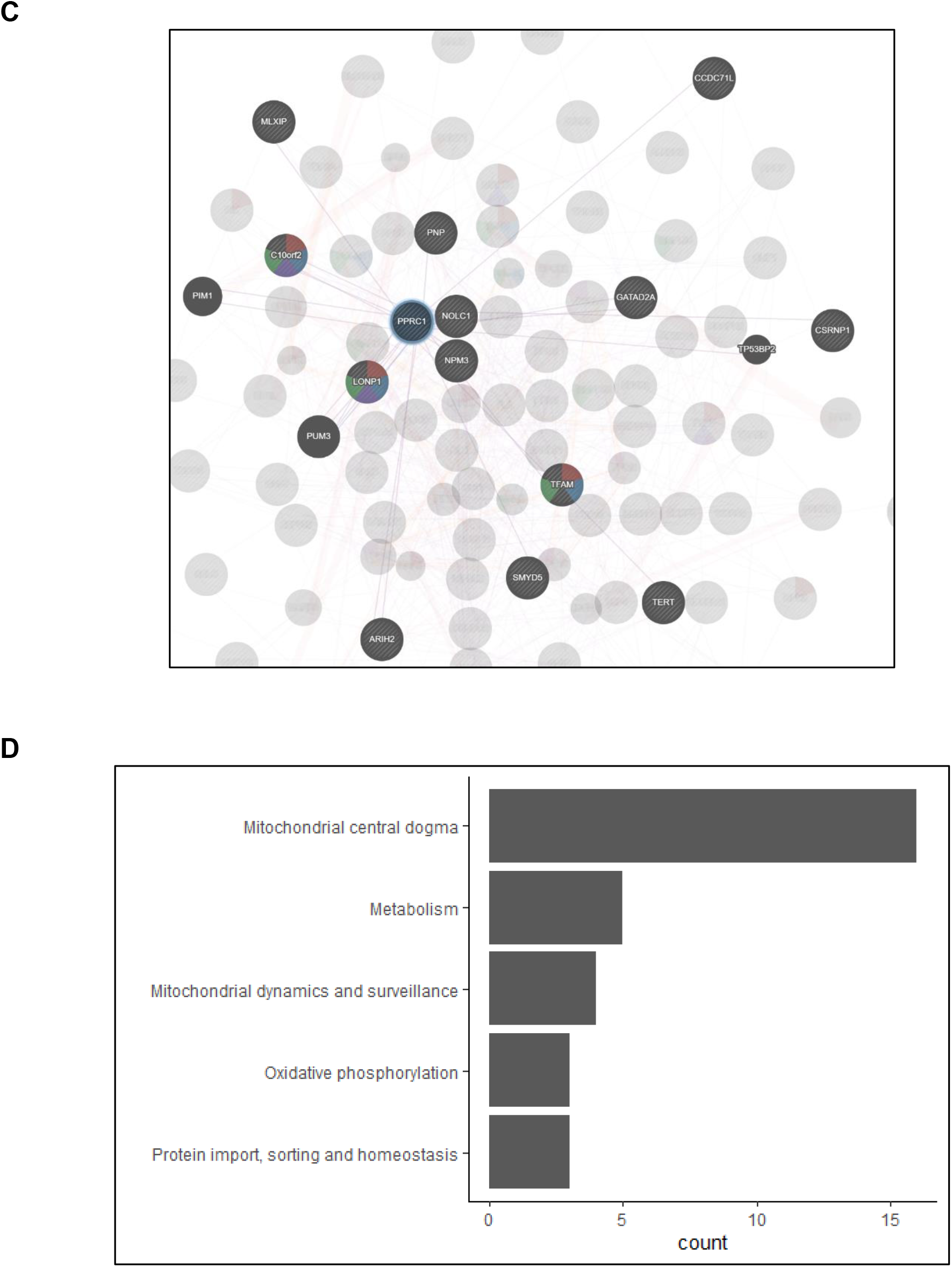
Analyses of common genetic loci associated with mtDNA-CN. (A) Manhattan plot illustrating common genetic variant associations with mtDNA-CN. (B) Size distribution of 95% credible sets defined for 82 independent genetic signals. (C) GENE-MANIA-mania protein network interaction exploration (D) “MitoPathway” counts corresponding to 27 prioritized MitoCarta3 genes encoding proteins with known mitochondrial localization.

**Figure 2.**
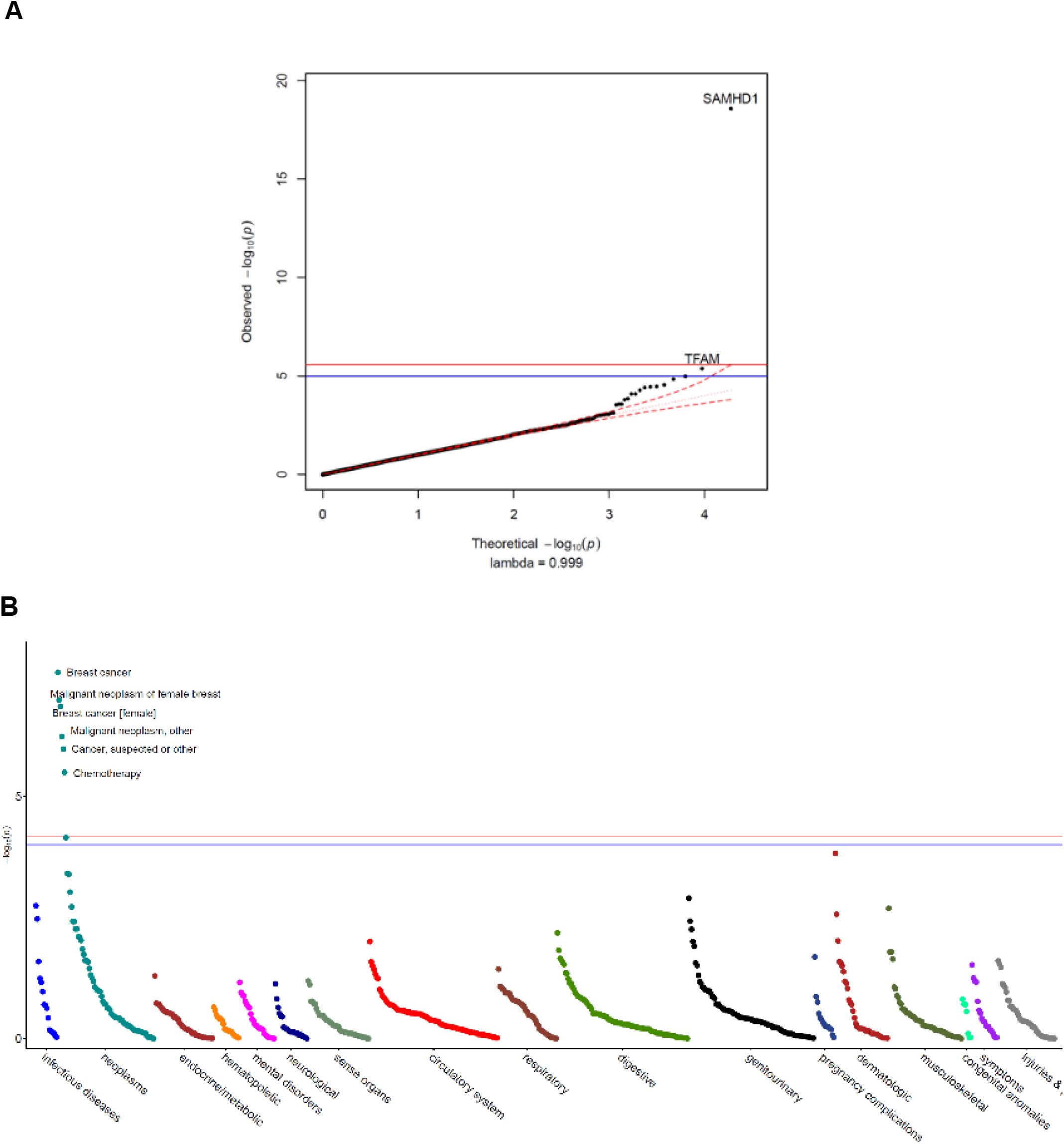
Rare variant gene burden association testing with mtDNA-CN and disease risk. (A) QQ plot illustrating expected vs. observed -log_10_(p) values for exome-wide burden of rare (MAF<0.001) and nonsynymous mutations. (B) Manhattan plot showing phenome-wide significant associations between *SAMHD1* carrier status and cancer-related phenotypes.

Fine-mapping via the FINEMAP algorithm (Benner et al. 2016) yielded 95% credible sets containing 2,363 genome-wide significant variants. Of the 82 independent genetic associations, 18 (22%) mapped to a single candidate causal variant; 32 (39%) mapped to 5 or fewer variants, and 42 (51%) mapped to 10 or fewer variants (Figure 1B; S2. Table 2). Credible sets for 11 genetic signals overlapped with genes responsible for rare mtDNA depletion disorders including *DGUOK* (3), *MGME1* (2), *TFAM* (2), *TWNK* (2), *POLG2* (1), and TYMP (1) (S2. Tables 3 & 4). Several associations mapped to coding variants with high posterior probability. *DGUOK* associations mapped to a synonymous variant (rs62641680; Posterior Probability=1) and a non-synonymous variant (rs74874677; PP=1). *TFAM* associations mapped to a 5’UTR variant (rs12247015; PP=1) falling within an ENCODE candidate cis-regulatory element with a promotor-like signature and an intronic variant (rs4397793; PP=1) with a proximal enhancer-like signature. Lastly, *POLG2* associations mapped to a nonsynonymous variant (rs17850455; PP=1). Beyond the six aforementioned mtDNA depletion genes identified at genome-wide significance, suggestive associations were found for *POLG* (rs2307441; P=1.0×10^−7^), *OPA1* (rs9872432; P=5.2×10^−7^), *SLC25A10* (rs62077224; P=1.2×10^−7^), and *RRM2B* (rs3907099; P=4.7×10^−6^). Given these observations, we hypothesized that mtDNA depletion genes may be generally enriched for common variant associations. Indeed, 10 (53%) of 19 known mtDNA depletion genes (Oyston 1998) harboured at least suggestive mtDNA-CN associations (P<5×10^−6^).

Additionally, trans-ethnic meta-analysis inclusive of non-Europeans (N=395,781) was performed but given the small increase in sample size, GWAS findings remained highly similar (S2. Figure 3). However, European effect estimates were significantly and highly correlated with those derived from South Asian (r=0.97; P= 2.2×10^−15^) and African (r=0.88; P=9.1×10^−5^) GWAS analyses (S2. Figure 4).

**Figure 3.**
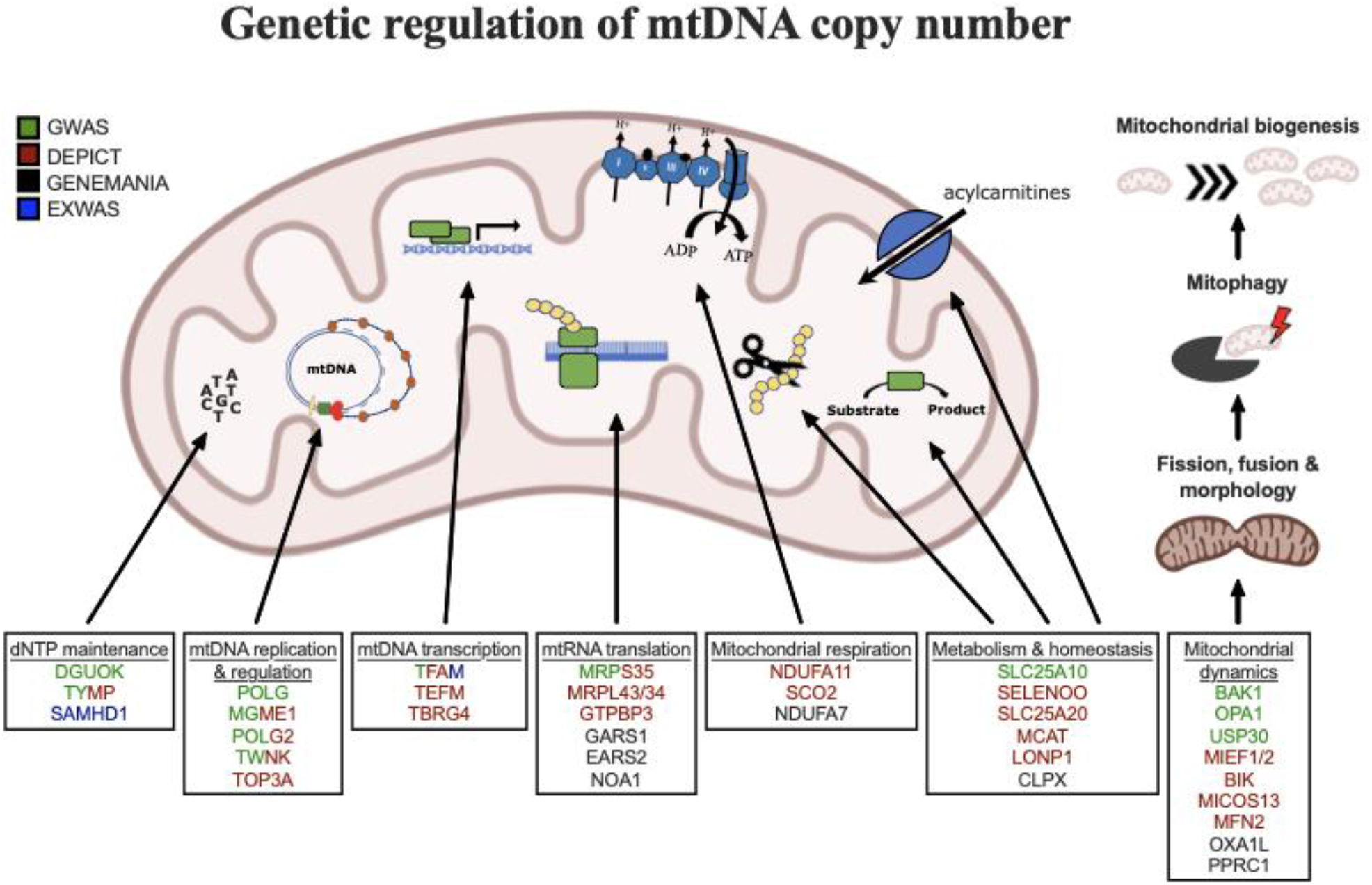
Graphical summary of mitochondrial genes and pathways implicated by genetic analyses. Colour-coding indicates through which set(s) of analyses genes were identified. The image was generated using BioRender (https://biorender.com/).

**Figure 4.**
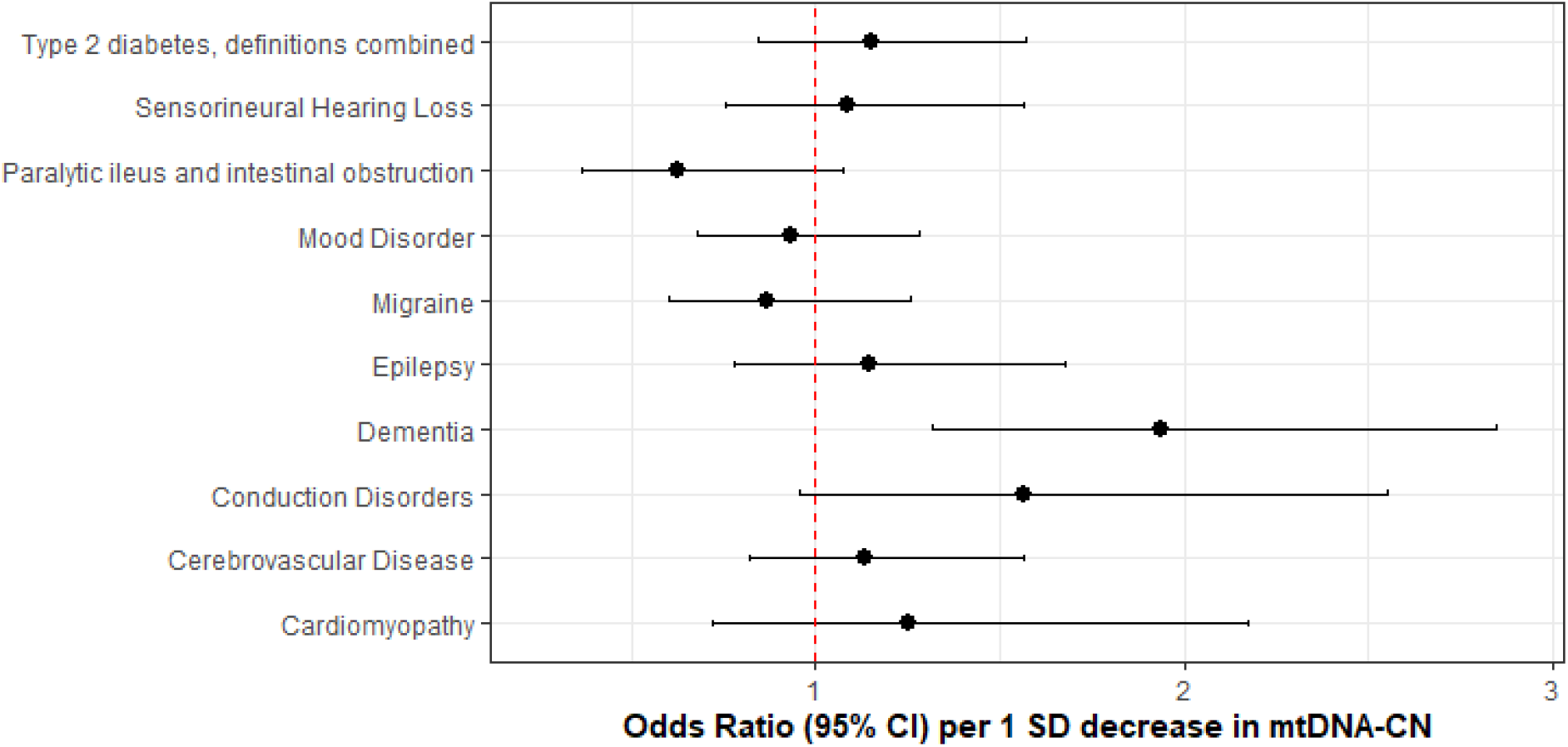
Coefficient plots for Mendelian Randomization analyses of mitochondrial disease traits. In the absence of heterogeneity (Egger-intercept P ≥ 0.05; MR-PRESSO global heterogeneity P ≥ 0.05), the inverse-variance weighted result was reported. In the presence of balanced pleiotropy (MR-PRESSO global heterogeneity P < 0.05), the weighted median result was reported. No set of analyses had evidence for directional pleiotropy (Egger-intercept P < 0.05).

### mtDNA-CN loci influence mitochondrial gene expression and heteroplasmy

We postulated that mtDNA-CN loci may regulate copy number by inducing changes in expression of genes that are directly transcribed from mtDNA. Ali *et al*. (2019) recently conducted a GWAS to identify nuclear genetic variants associated with variation in mtDNA-encoded gene expression (i.e. mt-eQTLs) (Ali et al. 2019). Nonsynonymous variants in *LONP1* (rs11085147) and *TBRG4* (rs2304693), as well as an intronic variant in *MRPS35* (rs1127787), were associated with changes in MT gene expression across various tissues (S2. Table 3). Nominally associated mtDNA-CN loci were also observed to influence MT gene expression including intronic variants in both *PNPT1* (rs62165226; mtDNA-CN P=5.5×10^−5^) and *LRPPRC* (rs10205130; mtDNA-CN P=1.1×10^−4^). Although differences in mitochondrial gene expression may be a consequence rather than a cause of variable mtDNA-CN, the analysis performed by Ali *et al* (2019) was corrected for factors associated with global changes in the mitochondrial transcriptome (Ali et al. 2019). Moreover, the direction of effect estimates between mtDNA-CN and mt-eQTLs varied depending on gene and tissue context. Altogether, such findings imply that some mtDNA-CN loci may regulate mtDNA-CN by influencing mitochondrial gene expression.

A recent GWAS by Nandakumar *et al* (2021) also reported 20 loci for mtDNA heteroplasmy. While full genome-wide summary statistics were not publicly available to systematically lookup potential effects of the 82 mtDNA-CN GWS variants on mtDNA heteroplasmy, we performed the reverse lookup of whether mtDNA heteroplasmy loci influenced mtDNA-CN. Of 19 matching variants between the GWAS, four heteroplasmy loci were also associated with mtDNA-CN at genome-wide significance including variants nearby or within *TINCR/LONP1* (rs12461806; mtDNA-CN GWAS P=7.5 ×10^−88^), *TWNK/MPRL43* (rs58678340; P=1.3×10^−39^), *TFAM* (rs1049432; P=1.5 ×10^−21^), and *PRKAB1* (rs11064881; P=2.6 ×10^−10^) genes. No additional mtDNA heteroplasmy loci were identified to influence mtDNA-CN when using the suggestive significance threshold.

### Genes and pathways implicated in the regulation of mtDNA-CN

DEPICT analysis led to the prioritization of 91 out of 18,922 genes (FDR P < 0.05; S2. Table 4). 87 of these genes intersected with the GeneMANIA database and were uploaded to the GeneMANIA platform to identify additional functionally related genes (Warde-Farley et al. 2010). GeneMANIA analysis discovered an additional 20 related genes (S2. Table 5). Among the 107 total genes prioritized by DEPICT or GeneMANIA (Figure 1C), mitochondrial functions were significantly enriched in gene ontology terms including mitochondrion organization (coverage: 12/225 genes; FDR P =7.4×10^−5^), mitochondrial nucleoid (6/34; FDR P=2.2×10^−4^), mitochondrial genome maintenance (4/10; FDR P=6.8×10^−4^), and mitochondrial matrix (11/257; FDR P=6.8×10^−4^). Visual inspection of the links between key genes involved in these functions highlights *PPRC1*, a member of the PGC-1A family of mitochondrial biogenesis activators (Richard C. Scarpulla 2011), as a potential coordinator of mtDNA-related processes (Figure 1C).

MitoCarta3 is a comprehensive and curated inventory of 1,136 human proteins (1,120 nuclear) known to localize to the mitochondria based on experiments of isolated mitochondria from 14 non-blood tissues (Rath et al. 2021). We leveraged this recently updated database, that was absent from GeneMANIA, to conduct a complementary set of targeted analyses focused on mitochondrial annotations (S2. Table 5). First, we hypothesized that prioritized genes would be generally enriched for genes encoding the mitochondrial proteome. Overall, 27 (25%) of 107 genes had evidence of mitochondrial localization corresponding to a 4.2-fold enrichment (null expectation = 5.9%; P=1.0×10^−10^). Next, given that *PPRC1*, an activator of mitochondrial biogenesis, was prioritized by DEPICT analyses and then linked to central mtDNA regulators in GeneMANIA, we postulated that prioritized genes may be enriched for downstream targets of PGC-1A. PGC-1A induction resulted in a higher mean fold-change among prioritized genes (beta=1.48; 95% CI, 0.60 to 2.37) as compared to any mitochondrial gene (beta=1.19; 95% CI, −0.76 to 3.13; t-test P=0.04). Finally, we categorized the 27 MitoCarta3 genes into their respective pathways. Most (16; 57%) genes were members of the “Mitochondrial central dogma” pathway but other implicated pathways included “Metabolism”, “Mitochondrial dynamics and surveillance”, “Oxidative phosphorylation”, and “Protein import, sorting and homeostasis” pathways (Figure 1D). Four proteins were annotated as part of multiple pathways including *TYMP/SCO2, GTPBP3, MIEF1*, and *OXA1L* (S2. Table 5).

### Exome-wide association testing uncovers rare coding *SAMHD1* mutations as a determinant of mtDNA-CN levels and breast cancer risk

We performed an exome-wide association study (EXWAS) in 147,740 UKBiobank participants with WES data to assess the contribution of rare coding variants. Among 18,890 genes tested, *SAMHD1* was the only gene reaching exome-wide significance (Figure 2A; S2 Table 8). The carrier prevalence of rare *SAMHD1* mutations was 0.75%, and on average, mutation carriers had higher mtDNA-CN than non-carriers (beta=0.23 SDs; 95% CI, 0.18-0.29; P=2.6×10^−19^; S2. Figure 5). Also, while none of the 19 known mtDNA depletion genes reached Bonferroni significance, a suggestive association was found for *TFAM* (beta=-0.33; 95% CI, −0.47 to −0.19; P=4.2×10^−6^).

To evaluate whether rare *SAMHD1* mutations also influenced disease risk, we conducted phenome-wide association testing of 771 diseases within the UKBiobank. At phenome-wide significance, *SAMHD1* mutation carrier status was associated with approximately two-fold increased risk of breast cancer (OR=1.91; 95% CI, 1.52-2.40; P=2.7×10^−8^), as well as greater risk of “cancer (suspected or other)” (OR=1.52; 95% CI, 1.28-1.80; P=1.1×10^−6^; Figure 2B; S2 Table 9). Exclusion of breast cancer cases attenuated but did not nullify the association with “cancer (suspected or other)” (OR=1.36; 95% CI, 1.10-1.67; P=0.004) suggesting that *SAMHD1* mutations may also increase risk of other cancers, as has been shown for colon cancer(Rentoft et al. 2019). To understand whether differences in mtDNA-CN levels between *SAMHD1* mutation carriers was a consequence of cancer diagnosis, we repeated association testing with mtDNA-CN excluding cancer patients. In this analysis, the association with mtDNA-CN levels was not attenuated (beta=0.26; 95% CI, 0.19-0.32; P=7.8×10^−15^) suggesting that the effect of rare *SAMHD1* variants on mtDNA-CN levels is not driven by its relationship with cancer status. A summary of mitochondrial genes and pathways implicated by common and rare loci is provided in Figure 3.

### Mendelian Randomization analysis implicates low mtDNA-CN as a causal mediator of dementia

Given that common variant loci overlapped with several mtDNA depletion genes, we postulated that polygenically low mtDNA-CN might cause a milder syndrome with phenotypically similar manifestations. To assess whether mtDNA-CN may represent a putative mediator of mtDNA depletion-related phenotypes, we conducted Mendelian Randomization analyses between genetically determined mtDNA-CN and mitochondrial disease phenotypes using summary statistics derived from the FinnGen v4 GWAS dataset.

After accounting for multiple testing of 10 phenotypes, an association between mtDNA-CN and all-cause dementia was found (OR=1.94 per 1 SD decrease in mtDNA-CN; 95% CI, 1.55-2.32; P=7.5×10^−4^; S2. Table 9; Figure 4). Sensitivity analyses indicated no evidence of global (MR-PRESSO P=0.51; Q-statistic P=0.51) or directional (Egger Intercept P =0.47) pleiotropy. The 27 selected variants accounted for 0.70% of the variance in mtDNA-CN and 0.13% of the risk for dementia, consistent with a causal effect of mtDNA-CN on dementia risk and not vice versa (Steiger P= 1.9×10^−62^). Findings were robust across several different MR methods including the Weighted Median (OR=2.47; 95% CI, 1.93-3.00; P=0.001) and MR-EGGER (OR=2.41; 95% CI, 1.71-3.11; P=0.02) methods. Furthermore, we replicated this result using a second UKBiobank-independent GWAS dataset derived from the International Genomics of Alzheimer’s Disease Consortium (2014) including 17,008 Alzheimer’s disease patients (OR=1.41; 95% CI, 1.0001-1.98; P=0.04993).

## Discussion

We developed a novel method to estimate mtDNA-CN from genetic array data, “AutoMitoC”, and applied it to the UKBiobank study. Extensive genetic investigations led to several key insights regarding mtDNA-CN. First, several novel common and rare genetic determinants of mtDNA-CN were identified, totalling 73 loci. Second, these loci were enriched for mitochondrial processes related to dNTP metabolism and the replication, packaging, and maintenance of mtDNA. Third, we observed a strong role for common variation within known mtDNA depletion genes in regulating mtDNA-CN in the general population. Fourth, we found that rare variants in *SAMHD1* not only affect mtDNA-CN levels but also confer risk to cancer. Finally, we provided the first Mendelian Randomization evidence implicating low mtDNA-CN as a causative risk factor for dementia.

While several investigations for mtDNA-CN have been performed, the present study represents the most comprehensive genetic assessment to date(Cai et al. 2015; Hägg et al. 2020; Ryan Joseph Longchamps 2019). Notably, Hagg *et al*. (2020) recently conducted a GWAS for mtDNA-CN in 295,150 UKBiobank participants and identified 50 common loci(Hägg et al. 2020). However, the method developed by Hagg *et al*. (2020) calibrated SNP probe intensities based on association with whole-exome sequencing read depths, which may limit the convenience of the method. In contrast, AutoMitoC only necessitates array probe intensities and does not require any secondary genetic measurements (WES or otherwise) for calibration. In addition, AutoMitoC exhibits superior concordance with WES-based estimates (Hagg r=0.33; AutoMitoC r=0.45), which was validated in an independent dataset with gold standard qPCR measurements. Further, Hagg *et al*. (2020) restricted genetic analyses to unrelated European individuals, whereas we incorporated ∼100,000 additional individuals and demonstrated consistency in genetic effects between Europeans and non-Europeans (r ≥ 0.88). The greater sample size in combination with more accurate mtDNA-CN estimates may explain the 44% increase in identified common loci (72 vs 50). Finally, in the present study we included exploration of the role of rare variants through ExWAS and, notably, Mendelian Randomization analyses to assess disease contexts whereby mtDNA-CN may represent a causal mediator and a potential therapeutic target.

MtDNA-CN has proven to be a biomarker of cardiovascular disease in several large epidemiological studies, with studies often assuming that such relationships are attributable to pathological processes including mitochondrial dysfunction, oxidative stress and inflammation (Fazzini et al. 2019; Koller et al. 2020; Tin et al. 2016; I.-C. Wu et al. 2017). Consistent with previous GWAS findings, our genetic analyses confirm that mtDNA-CN indeed reflects specific mitochondrial functions, but perhaps not the ones commonly attributable to mtDNA-CN(Cai et al. 2015; Guyatt et al. 2019; Hägg et al. 2020). Primarily, differences in mtDNA-CN reflect mitochondrial processes related to dNTP metabolism and the replication, maintenance, and organization of mtDNA. Secondarily, genes involved in mitochondrial biogenesis, metabolism, oxidative phosphorylation, and protein homeostasis were also identified but do not represent the main constituents. The observed enrichment in common variant associations within mtDNA depletion genes further reinforces the notion that differences in mtDNA-CN first and foremost reflect perturbations in mtDNA-related processes.

No therapy for mtDNA depletion disorders currently exists with treatment mainly consisting of supportive care. Intriguingly, we found that rare variants within *SAMHD1* were associated with increased levels of mtDNA-CN. *SAMHD1* is a multifaceted enzyme with various functions including tumour suppression through DNA repair activity and maintenance of steady-state intracellular dNTP levels, which has been involved in HIV-1 replication (Baldauf et al. 2012; Kretschmer et al. 2015). Rare homozygous and compound heterozygous loss-of-function mutations in *SAMHD1* result in an immune encephalopathy known as Aicardi Goutiere’s syndrome(White et al. 2017). Imbalanced intracellular dNTP pools and chronic DNA damage cause persistent elevations in interferon alpha thus mimicking a prolonged response to HIV-1 infection. While Aicardi Goutiere’s syndrome is a severe recessive genetic disorder, case reports of *SAMHD1*-related disease often describe heterozygous parents and siblings as being unaffected or with milder disease (familial chilblain lupus 2) (Haskell et al. 2018). In the UKBiobank, the vast majority (99.4%) of individuals possessing *SAMHD1* mutations were heterozygote carriers, who had a two-fold increased risk of breast cancer. Indeed, our finding that *SAMHD1* mutations associate with both elevated mtDNA-CN levels and risk of breast cancer belies the prevailing notion that higher mtDNA-CN is always a protective signature of proper mitochondrial function and healthy cells. Such findings may have important clinical implications for genetic screening. Firstly, heterozygous SAMHD1 mutations may be an overlooked risk factor for breast cancer considering that ∼1 in 130 UKBiobank participants possessed a genetic mutation conferring two-fold elevated risk. Notably, while *SAMHD1* mutations have been described previously to be associated with various cancers (Kohnken, Kodigepalli, and Wu 2015; Rentoft et al. 2019), this gene is not routinely screened nor part of targeted gene panels outside the context of neurological disorders (https://www.genedx.com/test-catalog/available-tests/comprehensive-common-cancer-panel/). Secondly, unaffected parents and siblings of Aicardi Goutiere patients might also present with greater risk of cancer. Thirdly, while SAMHD1 is a highly pleiotropic protein, therapeutic strategies to dampen (but not abolish) SAMHD1 activity might be considered to treat mtDNA depletion disorders caused by defects in nucleotide metabolism. Indeed, Franzolin *et al*. (2015) demonstrated that siRNA knockdown of SAMHD1 in human fibroblasts with *DGUOK* mtDNA depletion mutations partially recovered mtDNA-CN (Franzolin et al. 2015).

To our knowledge, we provide the first Mendelian Randomization evidence that mtDNA-CN may be causally related to risk of dementia. Although dysfunctional mitochondria have long been implicated in the pathogenesis of Alzheimer’s disease, only recently has mtDNA-CN been tested as a biomarker. Silzer *et al*. (2019) conducted a matched case-control study of 46 participants and showed that individuals with cognitive impairment had significantly lower blood-based mtDNA-CN(Silzer et al. 2019). Andrews *et al*. (2020) studied the relationship between post-mortem brain tissue mtDNA-CN and measures of cognitive impairment in 1,025 samples (Andrews and Goate 2020). Consistent with our findings, a 1 SD decrease in brain mtDNA-CN was associated with lower mini mental state exam (beta = −4.02; 95% CI, −5.49 to −2.55; P=1.07×10^−7^) and higher clinical dementia rating (beta = 0.71; 95% CI, 0.51 to 0.91). Both studies implicate blood and brain-based mtDNA-CN as a marker of dementia but were retrospective. In contrast, Yang *et al*. (2020) observed a significant association between mtDNA-CN and incident risk of neurodegenerative disease (Parkinson’s and Alzheimer’s disease)(Yang et al. 2021). Altogether, our results combined with previous findings suggest that mtDNA-CN represents both a marker and mediator of dementia. Considering that our overall findings suggest that mtDNA-CN reflects numerous mitochondrial subprocesses, future studies will be required to disentangle which ones, as reflected by diminished mtDNA-CN, truly mediate dementia pathogenesis.

Several limitations should be noted. First, mtDNA-CN approximated by array-based methods remain imperfectly accurate as compared to qPCR or whole genome sequencing measurements, though we found strong correlation between AutoMitoC and qPCR-based estimates in this study (r=0.64; P<2.23×10^−308^). Although whole genome sequencing will eventually supplant array-based mtDNA-CN GWAS, we hypothesize that the improvements made in the areas of speed, portability to ethnically diverse studies, and ease-of-implementation, should greatly increase accessibility of mtDNA-CN research as a plethora of genotyping array data is presently available to re-analyze. Third, Mendelian Randomization analyses were underpowered to conduct a broad survey of diseases in which mitochondrial dysfunction may play a causal role, and equally as important, we were unable to differentiate whether specific mitochondrial subpathways mediated risk of disease. As additional loci are uncovered, such analyses may be feasible. Fourth, variants and genes implicated in the regulation of mtDNA-CN may be specific to blood samples though findings suggest that many mtDNA-CN loci act through genes that are widely expressed in mitochondria across multiple tissues. Future studies are required to determine whether associations are ubiquitous across mitochondria-containing cells and to investigate the role of mtDNA-CN in other tissues. Lastly, whole blood mtDNA-CN reflects a heterogenous mixture of nucleated and unnucleated cells, and despite adjustment for major known confounding cell types, inter-individual differences in cell subpopulations not captured by a standard blood cell count may represent an important source of confounding.

## Conclusion

Although commonly viewed as a simple surrogate marker for the number of mitochondria present within a sample, genetic analyses suggest that mtDNA-CN is a highly complex biomarker under substantial nuclear genetic regulation. mtDNA-CN reflects a mixture of mitochondrial processes mostly pertaining to mtDNA regulation. Accordingly, the true relationship between mtDNA-CN measured in blood samples with human disease remains to be completely defined though we find evidence for mtDNA-CN as a putative causal risk factor for dementia. Future studies are necessary to decipher if mtDNA-CN is directly involved in the pathogenesis of dementia and other diseases or whether other specific mitochondrial processes are truly causative.

## Data Availability

Individual-level UKBiobank genotypes and phenotypes can be acquired upon successful application (https://bbams.ndph.ox.ac.uk/ams/). All individual-level UKBiobank data was accessed as part of application # 15255. FinnGen summary statistics are freely available to download (https://www.finngen.fi/en/access_results). All data products generated as part of this study will be made publicly accessible. Specifically, a software package for the AutoMitoC array-based mtDNA-CN estimation pipeline will be put on GitHub. The mtDNA-CN estimates derived in UKBiobank participants will be returned to the UKBiobank and made accessible to researchers through the data showcase (https://biobank.ndph.ox.ac.uk/showcase/). Summary-level association statistics from GWAS will be made publicly available for download. All remaining data are available in the main text or supplementary materials.

## Acknowledgements

We want to acknowledge the participants and investigators of the UKBiobank and FinnGen studies.

## Funding

Canadian Institutes of Health Research Frederick Banting and Charles Best Canada Graduate Scholarships Doctoral Award (MC, PM)

Canadian Institutes of Health Research Post-Doctoral Fellowship Award (RM)

Wellcome Trust Grant number: 099313/B/12/A; Crasnow Travel Scholarship; Bongani Mayosi UCT-PHRI Scholarship 2019/2020 (TM)

Wellcome Trust Health Research Board Irish Clinical Academic Training (ICAT) Programme Grant Number: 203930/B/16/Z (CJ)

European Research Council COSIP Grant Number: 640580 (MO)

E.J. Moran Campbell Internal Career Research Award (MP)

CISCO Professorship in Integrated Health Systems (GP)

Canada Research Chair in Genetic and Molecular Epidemiology (GP)

## Author Contributions

Conceptualization: MC, GP

Methodology: MC, WN, GP

Bioinformatics and Statistical Analysis: MC, PM, NP, GP, SN, IK, RL, MK, TM

Visualization: MC, PM, NP, RM, CJ

Data Interpretation: MC, PM, GP, SN, LA, RM

Funding or data acquisition: GP, MO, NC

Project administration: GP

Supervision: GP

Writing – original draft: MC

Writing – review & editing: MC, PM, NP, WN, RM, SN, IK, RL, MK, CJ, TM, NC, MO, MP, LA, GP

## Ethics Statement

Approval was received to use UKBiobank study data in this work under application ID # 15255 (“Identification of the shared biological and sociodemographic factors underlying cardiovascular disease and dementia risk”). The UKBiobank study obtained ethics approval from the North West Multi-centre Research Ethics Committee which encompasses the UK (REC reference: 11/NW/0382). All research participants provided informed consent.

## References and Notes

Abecasis, Goncalo R et al. 2012. “An Integrated Map of Genetic Variation from 1,092 Human Genomes.” Nature 491(7422): 56–65. http://www.pubmedcentral.nih.gov/articlerender.fcgi?artid=3498066&tool=pmcentrez&rendertype=abstract (July 9, 2014).

Ali, Aminah T. et al. 2019. “Nuclear Genetic Regulation of the Human Mitochondrial Transcriptome.” eLife 8: 1–23.

Andrews, Shea J, and Alison M Goate. 2020. “Mitochondrial DNA Copy Number Is Associated with Cognitive Impairment.” Alzheimer’s & Dementia 16(S5). https://onlinelibrary.wiley.com/doi/10.1002/alz.047543.

Ashar, Foram N et al. 2017. “Association of Mitochondrial DNA Copy Number With Cardiovascular Disease.” 21205(11): 1247–55.

Baldauf, Hanna-Mari et al. 2012. “SAMHD1 Restricts HIV-1 Infection in Resting CD4+ T Cells.” Nature Medicine 18(11): 1682–88. https://www.ncbi.nlm.nih.gov/pmc/articles/PMC3624763/pdf/nihms412728.pdf.

Benner, Christian et al. 2016. “FINEMAP: Efficient Variable Selection Using Summary Data from Genome-Wide Association Studies.” Bioinformatics 32(10): 1493–1501.

Benner, Christian et al. 2017. “Prospects of Fine-Mapping Trait-Associated Genomic Regions by Using Summary Statistics from Genome-Wide Association Studies.” The American Journal of Human Genetics 101(4): 539–51. https://linkinghub.elsevier.com/retrieve/pii/S0002929717303348.

Burbulla, Lena F. et al. 2017. “Dopamine Oxidation Mediates Mitochondrial and Lysosomal Dysfunction in Parkinson’s Disease.” Science 357(6357): 1255–61.

Cai, Na et al. 2015. “Genetic Control over MtDNA and Its Relationship to Major Depressive Disorder.” Current Biology 25(24): 3170–77. http://dx.doi.org/10.1016/j.cub.2015.10.065.

Denny, Joshua C et al. 2013. “Systematic Comparison of Phenome-Wide Association Study of Electronic Medical Record Data and Genome-Wide Association Study Data.” Nature biotechnology 31(12): 1102–10. http://www.ncbi.nlm.nih.gov/pubmed/24270849.

Desdín-micó, Gabriela et al. 2020. “T Cells with Dysfunctional Mitochondria Induce Multimorbidity and Premature Senescence.” 1376(June): 1371–76.

Fazzini, Federica et al. 2018. “Plasmid-Normalized Quantification of Relative Mitochondrial DNA Copy Number.” (May): 1–11.

Fazzini, Federica et al. 2019. “Mitochondrial DNA Copy Number Is Associated with Mortality and Infections in a Large Cohort of Patients with Chronic Kidney Disease.” 8: 480–88.

Franzolin, Elisa, Cristiano Salata, Vera Bianchi, and Chiara Rampazzo. 2015. “The Deoxynucleoside Triphosphate Triphosphohydrolase Activity of SAMHD1 Protein Contributes to the Mitochondrial DNA Depletion Associated with Genetic Deficiency of Deoxyguanosine Kinase.” Journal of Biological Chemistry 290(43): 25986–96.

Gene, The, and Ontology Consortium. 2000. “Gene Ontology?: Tool for The.” 25(may): 25–29.

Gorman Gráinne S. et al. 2016. “Mitochondrial Diseases.” Nature Reviews Disease Primers 2.

Guyatt, Anna L et al. 2019. “A Genome-Wide Association Study of Mitochondrial DNA Copy Number in Two Population-Based Cohorts.” : 1–17.

Hägg, Sara et al. 2020. “Deciphering the Genetic and Epidemiological Landscape of Mitochondrial DNA Abundance.” Human Genetics (0123456789). https://doi.org/10.1007/s00439-020-02249-w.

Haskell, Gloria T. et al. 2018. “Combination of Exome Sequencing and Immune Testing Confirms Aicardi–Goutières Syndrome Type 5 in a Challenging Pediatric Neurology Case.” Molecular Case Studies 4(5): a002758. http://molecularcasestudies.cshlp.org/lookup/doi/10.1101/mcs.a002758.

Hemani, Gibran et al. 2018. “The MR-Base Platform Supports Systematic Causal Inference across the Human Phenome.” eLife 7: 1–29.

Hu, Liwen, Xinyue Yao, and Yi Shen. 2016. “Altered Mitochondrial DNA Copy Number Contributes to Human Cancer Risk: Evidence from an Updated Meta-Analysis.” Scientific Reports 6(October): 1–11.

Jagadeesh, Karthik A et al. 2016. “M-CAP Eliminates a Majority of Variants of Uncertain Significance in Clinical Exomes at High Sensitivity.” Nature genetics 48(12): 1581– 86. http://www.ncbi.nlm.nih.gov/pubmed/27776117.

Kamat, Mihir A et al. 2019. “PhenoScanner V2: An Expanded Tool for Searching Human Genotype-Phenotype Associations.” Bioinformatics (Oxford, England) 35(22): 4851–53. http://www.ncbi.nlm.nih.gov/pubmed/31233103.

Kim, Christopher et al. 2015. “Mitochondrial DNA Copy Number and Chronic Lymphocytic Leukemia/Small Lymphocytic Lymphoma Risk in Two Prospective Studies.” Cancer Epidemiology Biomarkers and Prevention 24(1): 148–53.

Kohnken, Rebecca, Karthik M. Kodigepalli, and Li Wu. 2015. “Regulation of Deoxynucleotide Metabolism in Cancer: Novel Mechanisms and Therapeutic Implications.” Molecular Cancer 14(1): 1–11.

Koller, A. et al. 2020. “Mitochondrial DNA Copy Number Is Associated with All-Cause Mortality and Cardiovascular Events in Patients with Peripheral Arterial Disease.” Journal of Internal Medicine 287(5): 569–79.

Kretschmer, Stefanie et al. 2015. “SAMHD1 Prevents Autoimmunity by Maintaining Genome Stability.” Annals of the Rheumatic Diseases 74(3).

Lane, John. 2014. “MitoPipeline: Generating Mitochondrial Copy Number Estimates from SNP Array Data in Genvisis.” http://genvisis.org/MitoPipeline/.

Longchamps, Ryan J. et al. 2020. “Evaluation of Mitochondrial DNA Copy Number Estimation Techniques” ed. David C. Samuels. PLOS ONE 15(1): e0228166. https://dx.plos.org/10.1371/journal.pone.0228166.

Longchamps, Ryan Joseph. 2019. “EXPLORING THE ROLE OF MITOCHONDRIAL DNA QUANTITY AND QUALITY.” Biorxiv (August).

Madewell, Zachary J et al. 2020. “Findings and Insights from the Genetic Investigation of Age of First Reported Occurrence for Complex Disorders in the UK Biobank and FinnGen.” medRxiv: 1–13.

Mbatchou, Joelle et al. 2020. “Computationally Efficient Whole Genome Regression for Quantitative and Binary Traits.” : 1–88.

Nandakumar, Priyanka et al. 2021. “Nuclear Genome-Wide Associations with Mitochondrial Heteroplasmy.” Science Advances 7(12): 1–10.

Oyston, J. 1998. “Online Mendelian Inheritance in Man.” Anesthesiology 89(3): 811–12.

Pers, Tune H. et al. 2015. “Biological Interpretation of Genome-Wide Association Studies Using Predicted Gene Functions.” Nature Communications 6: 5890. http://www.pubmedcentral.nih.gov/articlerender.fcgi?artid=4420238&tool=pmcentrez&rendertype=abstract.

Purcell, Shaun et al. 2007. “PLINK: A Tool Set for Whole-Genome Association and Population-Based Linkage Analyses.” American journal of human genetics 81(3): 559–75.

Rath, Sneha et al. 2021. “MitoCarta3.0: An Updated Mitochondrial Proteome Now with Sub-Organelle Localization and Pathway Annotations.” Nucleic acids research 49(D1): D1541–47.

Rentoft, Matilda et al. 2019. “Erratum: Heterozygous Colon Cancer-Associated Mutations of SAMHD1 Have Functional Significance (Proceedings of the National Academy of Sciences of the United States of America (2016) 113 (4723-4728) DOI: 10.1073/Pnas.1519128113).” Proceedings of the National Academy of Sciences of the United States of America 116(10): 4744.

Richard C. Scarpulla. 2011. “Metabolic Control of Mitochondrial Biogenesis through the PGC-1 Family Regulatory Network.” Biochim Biophys Acta. 1813(7): 1269–78.

Silzer, Talisa et al. 2019. “Circulating Mitochondrial DNA?: New Indices of Type 2 Diabetes-Related Cognitive Impairment in Mexican Americans.” : 1–21.

Simone, Domenico et al. 2011. “The Reference Human Nuclear Mitochondrial Sequences Compilation Validated and Implemented on the UCSC Genome Browser.” BMC genomics 12: 517. http://www.ncbi.nlm.nih.gov/pubmed/22013967.

Sliter, Danielle A et al. 2020. “Parkin and PINK1 Mitigate STING-Induced Inflammation.” Nature 561(7722): 258–62.

Sudlow, Cathie et al. 2015. “UK Biobank: An Open Access Resource for Identifying the Causes of a Wide Range of Complex Diseases of Middle and Old Age.” PLoS medicine 12(3): e1001779. http://www.ncbi.nlm.nih.gov/pubmed/25826379.

Tin, Adrienne et al. 2016. “Association between Mitochondrial DNA Copy Number in Peripheral Blood and Incident CKD in the Atherosclerosis Risk in Communities Study.” Journal of the American Society of Nephrology 27(8): 2467–73.

Verbanck, Marie, Chia Yen Chen, Benjamin Neale, and Ron Do. 2018. “Detection of Widespread Horizontal Pleiotropy in Causal Relationships Inferred from Mendelian Randomization between Complex Traits and Diseases.” Nature Genetics 50(5): 693–98. http://dx.doi.org/10.1038/s41588-018-0099-7.

Vuckovic, Dragana et al. 2020. “The Polygenic and Monogenic Basis of Blood Traits and Diseases.” Cell 182(5): 1214-1231.e11. http://www.ncbi.nlm.nih.gov/pubmed/32888494.

Warde-Farley, David et al. 2010. “The GeneMANIA Prediction Server: Biological Network Integration for Gene Prioritization and Predicting Gene Function.” Nucleic Acids Research 38(SUPPL. 2): 214–20.

Wei, Wei Qi et al. 2017. “Evaluating Phecodes, Clinical Classification Software, and ICD-9-CM Codes for Phenome-Wide Association Studies in the Electronic Health Record.” PLoS ONE 12(7): 1–16.

White, Tommy E. et al. 2017. “A SAMHD1 Mutation Associated with Aicardi-Goutières Syndrome Uncouples the Ability of SAMHD1 to Restrict HIV-1 from Its Ability to Downmodulate Type I Interferon in Humans.” Human Mutation 38(6): 658–68. http://doi.wiley.com/10.1002/humu.23201.

Willer, Cristen J., Yun Li, and Gonçalo R. Abecasis. 2010. “METAL: Fast and Efficient Meta-Analysis of Genomewide Association Scans.” Bioinformatics 26(17): 2190– 91.

Wu, I-Chien et al. 2017. “Interrelations Between Mitochondrial DNA Copy Number and Inflammation in Older Adults.” The Journals of Gerontology: Series A 72(7): 937– 44. https://academic.oup.com/biomedgerontology/article/72/7/937/3065927.

Wu, Patrick et al. 2019. “Mapping ICD-10 and ICD-10-CM Codes to Phecodes: Workflow Development and Initial Evaluation.” JMIR medical informatics 7(4): e14325. http://www.ncbi.nlm.nih.gov/pubmed/31553307.

Yang, Stephanie Y. et al. 2021. “Blood-Derived Mitochondrial DNA Copy Number Is Associated with Gene Expression across Multiple Tissues and Is Predictive for Incident Neurodegenerative Disease.” Genome Research. http://genome.cshlp.org/lookup/doi/10.1101/gr.269381.120.

Zhang, Yiyi et al. 2017. “Association between Mitochondrial DNA Copy Number and Sudden Cardiac Death?: Findings from the Atherosclerosis Risk in Communities Study (ARIC).” : 3443–48.

