## Supplementary Material for "GWAS and ExWAS of blood Mitochondrial DNA copy number identifies 73 loci and highlights a potential causal role in dementia"

### Supplementary Material and Methods

#### Supplementary Results I:
#### Development of the Automatic Mitochondrial Copy (AutoMitoC) Number Pipeline

##### Background & Premise Underlying Array-based mtDNA-CN Estimation

While SNP array data is intended for highly multiplexed determination of genotypes, the raw probe signal intensities used for genotypic inference can also be co-opted to derive estimates of mtDNA-CN. Determining genotypes for a sample at a given variant site relies on contrasting hybridization intensities of allele-specific oligonucleotide probes and then assigning membership to the most probable genotype cluster based on intensity properties (S1. Figure 1). Variation in intensities within each genotyping cluster can also be co-opted to deduce variations in copy number. A commonly used metric of probe intensity is the "log2ratio" (L2R), which denotes $\log_2$(observed intensity / expected intensity), where the expected intensity is defined as the median signal intensity for a probe conditional on each genotype cluster.

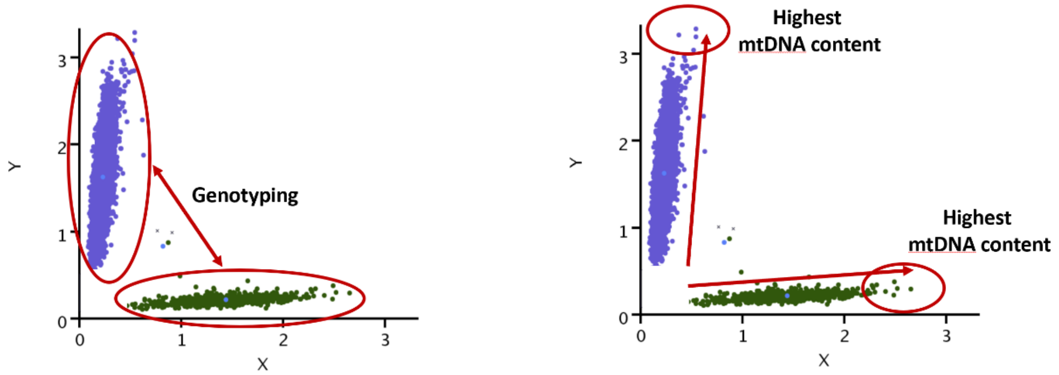


**S1. Figure 1.** Contrast in the intensities of mitochondrial probes X and Y discriminate genotypes. Intra-cluster variation in signal intensities may reflect mtDNA-CN. (Adapted from Lane *et al.*(Lane 2014))

##### Existing Methodology: The MitoPipeline

The "MitoPipeline" is a framework for estimating mtDNA-CN from array-based L2R (aka LRR) values developed by Lane *et al.* (2015) (Lane 2014; Zhang et al. 2017). An overview of the MitoPipeline is described in S1. Figure 2. To briefly summarize: (i) autosomal and mitochondrial L2R values are first corrected for GC waves; (ii) a high-quality set of mitochondrial and autosomal markers are selected largely based on visual inspection of genotype clusters and BLAST alignment for non-homologous sequences; (iii) principal component analysis (PCA) of at least 40,000 autosomal markers is conducted to capture batch effects; (iv) finally, mtDNA-CN is estimated based on median MT L2R value for each sample and then corrected for background noise through residualization of top autosomal PCs.

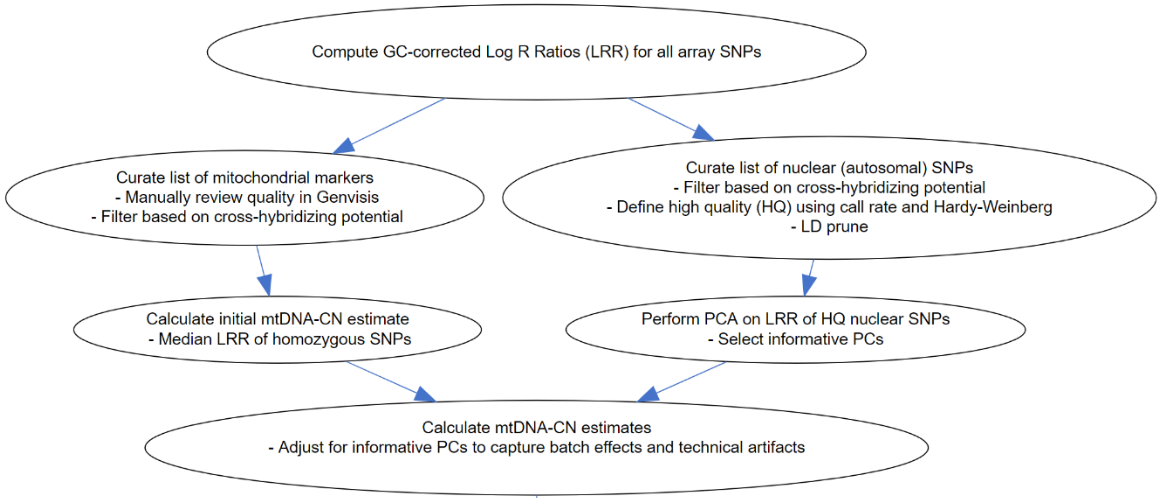


**S1. Figure 2.** Overview of the MitoPipeline (Source: http://genvisis.org/MitoPipeline/) (Lane 2014).

The MitoPipeline has proven to be effective in estimating mtDNA-CN (correlation coefficient R ~ 0.5 with direct qPCR estimates) as evidenced by multiple epidemiological studies employing this method (Ashar et al. 2017; Fazzini et al. 2019; Zhang et al. 2017). Firstly, visual inspection of MT probe intensity clusters is recommended to remove probes with poorly differentiated genotype clusters. However, this process is time-consuming (especially for biobank studies that are genotyped across thousands of batches); determination of probes with “good” vs “bad” genotype clustering is subjective; and guidance is only provided for adjudication of polymorphic but not monomorphic markers which may still be informative. Secondly, in consideration of nuclear and mitochondrial sequences with significant sequence similarity due to past and recurrent transposition of mitochondrial sequence into the nuclear genome, also known as “NUMTs” (Simone et al. 2011), the MitoPipeline recommends exclusion of MT probes with greater than 80% sequence similarity to the nuclear genome. While determining sequence homology of probes to the nuclear genome may have been feasible with older microarrays wherein probe sequences were often publicized, for many contemporary arrays, including the UKBiobank array, such information is not readily available. Thirdly, LD-pruning of common autosomal variants is required to ascertain a set of independent genetic variants, but implementation of this approach within ethnically diverse studies becomes more complex since genetic independence is ancestry-dependent. Under the MitoPipeline framework, each ethnicity warrants a unique set of common variants, which not only adds to computational burden but also creates an additional source of variability in terms of performance of the method between ethnicities.

#### Proposed Methodology: AutoMitoC

Therefore, we developed a new array-based mtDNA-CN estimation method, which we have dubbed the “AutoMitoC” pipeline, which incorporates three key amendments: (i) Autosomal signal normalization utilises globally rare variants in place of common variants which confers advantages in terms of both speed and portability to ethnically diverse studies, (ii) cross-hybridizing probes are identified by assessing evidence for cross-hybridization via association of signal intensities (rather than using genotype association and identification of homologous sequences through BLAST alignment), and (iii) the primary estimate of mitochondrial (MT) signal is ascertained using PCA as opposed to using the median signal intensity of MT probes. The rationale underlying these amendments are described in detail in the subsequent sections. An overview of the AutoMitoC pipeline is provided in S1. Figure 3.

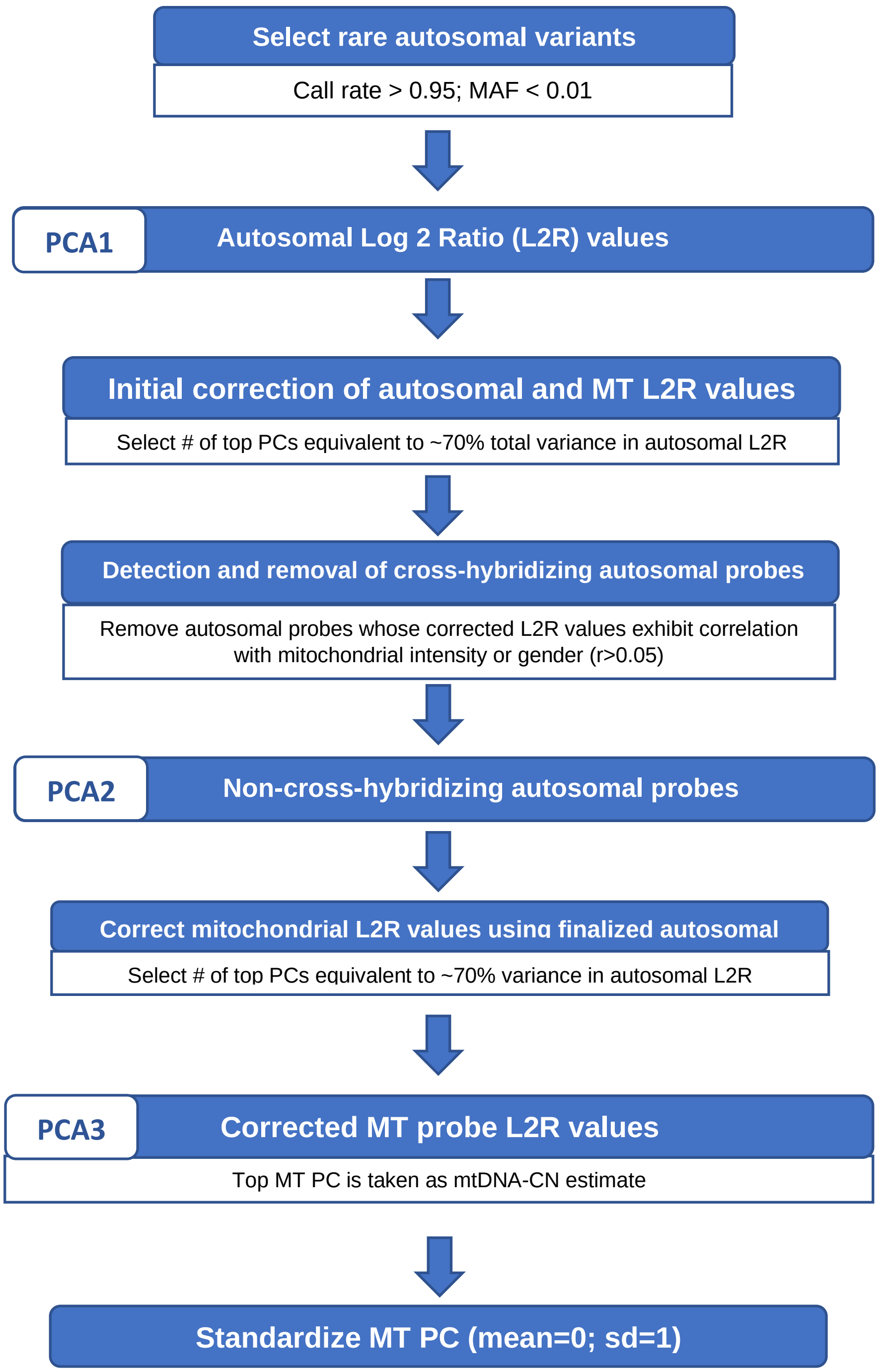


**S1. Figure 3.** Overview of the AutoMitoC Pipeline.

#### Development of AutoMitoC in the UKBiobank study

To develop the AutoMitoC pipeline we used genetic datasets from the large UKBiobank prospective cohort studywhich includes approximately half of a million UK residents recruited from 2006 to 2010 in whom extensive genotypic and phenotypic investigations have been and continue to be performed (Sudlow et al. 2015). The size, breadth, and depth of such investigations makes this a rich resource for both methodological development and medical research. All UKBiobank data was accessed as part of application ID: 15255, “Identification of the shared biological and sociodemographic factors underlying cardiovascular disease and dementia risk”. Two main genetic datasets from the UKBiobank were incorporated in the development of AutoMitoC. Firstly, CNV log2r (L2R) values derived from genetic arrays (i.e. normalized array probe intensities) for 488,264 samples were downloaded using the *ukbgene* utility, and their corresponding genotype calls (data field: 22418) were downloaded with *gfetch*. Secondly, exome alignment maps (EXOME FE CRAM files and indices; data fields 23163 & 23164) from the first tranche 49,989 samples released in March, 2019 were downloaded with the “ukbfetch” utility. L2R values were used to derive AutoMitoC mtDNA-CN estimates. For whole-exome sequencing data, Samtools *idxstats* was used to derive the number of sequence reads aligning to mitochondrial and autosomal genomes, from which an estimate of mtDNA-CN was derived according to the procedure by Longchamps *et al.* (2019) (Longchamps 2019); these complementary WES-based mtDNA-CN estimates served as a comparator to benchmark AutoMitoC performance.

Initial quality control of 488,264 samples and 784,256 directly genotyped variants was executed in PLINK following that of the Mitopipeline (i.e. sample call rate > 0.96; variant call rate > 0.98; HWE p-value > $1x10^{-5}$; PLINK mishap P-value > $1x10^{-4}$; genotype association with sex p-value > 0.00001; LD-pruning r2 < 0.30; MAF > 0.01) (Purcell et al. 2007). Variants within 1 Mb of immunoglobulin, T-cell receptor genes, and centromeric regions were removed. After this quality control procedure, 466,093 samples and 86,677 common variants remained. Next, genomic waves were corrected according to Diskin *et al.* (2008) using the PennCNV “genomic_wave.pl” script (https://github.com/WGLab/PennCNV/blob/master/genomic_wave.pl) (Diskin et al. 2008; Wang et al. 2007). Samples with high genomic waviness (L2R SD > 0.35) before and after GC-correction were removed resulting in 431,501 samples with array L2R values corresponding to 86,677 common autosomal variants. Lastly, we excluded samples representing blood cell count outliers as per Longchamps *et al.* (2019) which led to 395,781 participants (Longchamps 2019). Finally, we took the intersection of European samples with both suitable array and whole-exome sequencing data resulting in a final testing dataset of 34,436 European participants. To evaluate the possibility of replacing common autosomal variant signal normalization with rare variants, we also analyzed a set of 79,611 variants with a MAF$\leq$0.01.

#### Background correlation between autosomal & MT signal intensities

Inference of relative mitochondrial DNA copy number (mtDNA-CN) from array data consists of determining the ratio of mitochondrial to autosomal probe signal intensities (or normalized L2R values) within each sample. Technical (e.g. batch and plate effects) and latent sample factors confound raw signal intensities for reasons unrelated to DNA quantity. Such confounders induce strong cross-genome correlation, and therefore, a necessary first step is to remove this background noise from autosomal and mitochondrial probe intensities.

Indeed, even after correcting autosomal L2R values for genomic waves, we observed significant correlation between individual autosomal probe intensities and the median sample intensity across mitochondrial probes (S1. Figure 4A). The extent of cross-genome intensity correlation varied based on minor allele frequency (MAF), with rare autosomal variants (MAF<0.01; M=79,611) showing the strongest correlation. We postulate that intensity properties for rare variants, which have a higher prevalence of homozygous genotypes, more strongly resemble those for mitochondrial genotypes, which are predominantly homoplasmic. On this basis, we only use rare autosomal variants to represent autosomal signal. While this approach contrasts with the Mitopipeline, which utilises common genetic variants to represent autosomal signal, restricting autosomal signal normalization to rare variants confers three major advantages while maintaining the same level of concordance with WES estimates ($R_{common}$=0.50 ; $R_{rare}$=0.49). First, this allows for further streamlining of the pipeline as this precludes the necessity for common variant filters, such as Hardy Weinberg equilibrium or LD-pruning. Second, we show that fewer principal components (PCs) are necessary to capture the same proportion of total variance in signal intensities with rare as opposed to common variants. Approximately 70% of the total variance in rare autosomal intensities was explained by 120 PCs, whereas the same proportion of variance in common autosomal intensities would necessitate more than 1000 PCs (S1. Figure 4B). In the UKBiobank, PCs were derived via the eigendecomposition of the empirical covariance matrix conducted in Python 3.6, using NumPy and SciPy (Harris et al. 2020; Virtanen et al. 2020).

Third, the set of autosomal markers used in deriving mtDNA-CN remains independent from the set of common autosomal variants analyzed in subsequent GWAS for mtDNA-CN. Effectively, this ensures that common autosomal variants evaluated for association with mtDNA-CN in downstream GWAS analyses are not directly incorporated into autosomal signal normalization, which could otherwise attenuate GWAS signals.

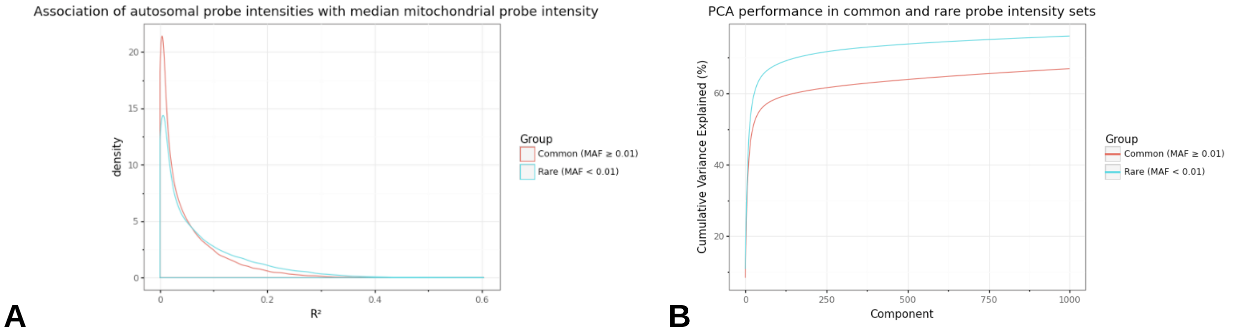


**S1. Figure 4**. (A) Histogram illustrating the square of the Pearson correlation coefficient ($R^2$) of autosomal GC-corrected L2R values with median mitochondrial signal intensity stratified by MAF categories. (B) Cumulative variance explained by inclusion of top eigenvectors for sets of common (MAF>0.01; M=86,677) and rare (MAF≤0.01; M=79,611) autosomal probe sets.

#### Empirical Detection of off-target probes

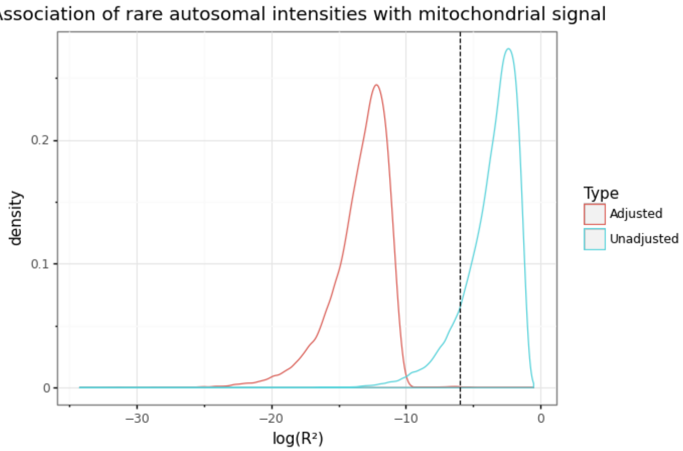


**S1. Figure 5**. Distribution of log10 transformed coefficients of determination ($R^2$) from the association between autosomal probe intensities vs. median mitochondrial signal with (blue) or without (red) correction for background noise (i.e. 120 autosomal PCs). The dashed vertical line represents the threshold corresponding to “moderate” correlation ($|R|>0.05$ or $R^2>0.0025$), which is used to remove outlying probes that are associated with mitochondrial signal. Without correction for top PCs, the most autosomal probes exhibit some correlation with mitochondrial signal.

After adjustment for 120 autosomal PCs (approximating the elbow of the variance explained curve), there persisted a smaller subset of autosomal probes that were significantly correlated with median MT intensity (S1. Figure 5). We hypothesized that such probes either (i) cross-hybridize with the MT genome (i.e. lie within nuclear mitochondrial DNA (NUMT) regions) or (ii) corresponded to genetic loci involved in regulation of mtDNA-CN. As an illustration, S1. Table 1 conveys characteristics of the 10 most strongly correlated variants. Four of the top 10 probes were located within 1Mb of a NUMT region, and in all four cases, the sign of the correlation coefficient was negative which might reflect interference of autosomal signal with increasing mtDNA-CN. An additional 3 probes corresponded to variants within genes that were implicated in mitochondrial disorders or regulation of mitochondrial processes (S1. Table 1).

**S1. Table 1**. Characteristics of top 10 autosomal probes whose adjusted intensities correlate with median mitochondrial signal.

| **Autosomal Probe ID** | **Genomic Coordinates** | **Intensity R (Auto vs. Mito)** | **Comment** |
|---|---|---|---|
| rs68130461 | 17:22024892 | -0.20 | HSA_NumtS_508_b2 |
| Affx-80229644 | 6:43484921 | -0.11 | HSA_NumtS_239_b1 (+25 Kb) |

| | | | |
|---|---|---|---|
| rs35201453 | 14:22783111 | -0.09 | NA |
| rs41267813 | 6:160998199 | -0.08 | HSA_NumtS_261_b1 (-742 Kb) |
| rs117507044 | 13:19958310 | -0.08 | NA |
| rs113200742 | 6:25272561 | -0.08 | HSA_NumtS_236_b1 (-324 Kb) |
| rs201397731 | 1:161168270 | 0.08 | *NDUFS2* Coding Variant (gene causes Mitochondrial Complex I Deficiency) |
| rs138167117 | 17:65739626 | 0.08 | NA |
| rs138656762 | 19:36330320 | 0.08 | *NPHS1* pathogenic variant (causes Finnish Nephrotic syndrome, characterized by mitochondrial dysfunction in kidneys) |
| rs117116233 | 19:8535980 | -0.08 | *HNRNPM* intronic variant (putative regulator of mitochondrial processes) |

We further explored whether there was evidence for cross-hybridization between autosomal SNPs and sex chromosomes by regressing adjusted autosomal probe intensities with reported male status. Generally, correlations between autosomal intensities and sex were stronger than those with MT intensities, suggesting that cross-hybridization of autosomal probes to sex chromosomes is more pronounced. For the top 10 sex-associated probes, we performed BLASTn alignment against the human reference genome (GRCh38) using 30 bases surrounding each probe (S1. Table 2) (Altschul et al. 1990). All probes had at least one flanking sequence with near-perfect ($\geq$ 97%) sequence identity to a sex chromosome. For these 10 probes, the sign of the correlation between autosomal intensity and male status was perfectly consistent with homology to X or Y chromosomes, thus supporting the hypothesis that such probes cross-hybridized to sex chromosomes.

**S1. Table 2.** Characteristics of top autosomal probes associated with male sex status.

| Autosomal Probe ID | Genomic Coordinates | Intensity R (Auto vs. Male Sex) | Flank (30 BP) | Chromosome | Sequence Similarity (E-value) |
|---|---|---|---|---|---|
| Affx-89012246 | 7:141336763 | 0.67 | Right | Y | 97% (9x10$^{-6}$) |
| rs138167117 | 17:65739626 | -0.60 | Left | X | 100% (3x10$^{-8}$) |
| rs147585440 | 18:47310224 | -0.60 | Left | X | 100% (3x10$^{-8}$) |
| Affx-80264600 | 21:38555134 | -0.59 | Left | X | 100% (3x10$^{-8}$) |
| Affx-80229637 | 6:43470088 | -0.59 | Left | X | 100% (3x10$^{-8}$) |
| rs56275071 | 10:88822514 | -0.57 | Left | X | 97% (9x10$^{-6}$) |
| rs117507044 | 13:19958310 | 0.57 | Left | Y | 100% (3x10$^{-8}$) |

| | | | | | |
|---|---|---|---|---|---|
| Affx-89008518 | 7:140501303 | -0.57 | Left | X | 100% ($3x10^{-8}$) |
| Affx-80224967 | 4:84380892 | -0.57 | Right | X | 100% ($3x10^{-8}$) |
| Affx-89005068 | 7:140501288 | -0.55 | Left | X | 100% ($9x10^{-6}$) |

Inclusion of autosomal probes with evidence of off-target hybridization to the mitochondrial genome or sex chromosomes is problematic. In the former scenario, misattribution of autosomal as mitochondrial signal may reduce the effectiveness of normalization. In the latter scenario, inadvertent adjustment for sex through retention of cross-hybridizing autosomal probes may occur if such probes explain substantial variance in autosomal probe intensities and this is particularly problematic given that mtDNA-CN has been robustly shown to differ between genders in epidemiological studies. In preliminary investigations where sex-associated probes were retained, we noticed that several top PCs that perfectly tagged gender. Hypothetically, had these PCs been retained and used for correction of mitochondrial signal, then the final mtDNA-CN estimate would have been inadvertently corrected for gender. Therefore, we removed autosomal probes exhibiting moderate correlation (|R| >0.05) with sex (907; 1.14%) or median mitochondrial intensity (193; 0.24%) and then recalculated top autosomal PCs.

**PCA-based approach improves concordance with complementary estimates**

After correcting MT probes using the updated set of 120 autosomal L2R PCs, we adopted the Mitopipeline's approach for estimating mtDNA-CN and calculated the median of corrected MT L2R values to denote an individual's final mtDN A-CN estimate. Using this median-based approach, array mtDNA-CN estimates demonstrated significant correlation with WES mtDNA-CN estimates ($R=0.33$; $P < 2.23x10^{-308}$). However, we found that performing PCA across all corrected MT L2R values and then extracting the top MT PC for each sample as the final mtDNA-CN estimate resulted in stronger correlation with WES ($R=0.49$; $P < 2.23x10^{-308}$).

**Independent validation in an ethnically diverse sample**

We additionally validated the AutoMitoC pipeline by deriving array-based mtDNA-CN estimates in the INTERSTROKE study and comparing these with parallel qPCR-based measurements, the current gold standard for measuring mtDNA-CN. INTERSTROKE is an international case-control study of stroke including 26,526 participants from 32 countries and 142 centers(O'Donnell et al. 2010). Blood samples have been collected for a subset of approximately 12,000 individuals, of which 9,311 have been successfully genotyped using the Axiom Precision Medicine Research Array (PMRA r3). A further subset of 5,791 samples with both suitable array genotypes have undergone qPCR measurement of mtDNA-CN using the plasmid-normalized protocol from Fazzini *et al.* (2019). Within INTERSTROKE, concordant findings to UKB-based analyses were observed favouring the PC-based ($r=0.64$; $P< 2.23x10^{-308}$) over the median-based approach (R=0.60; $P < 2.23x10^{-308}$). Furthermore, INTERSTROKE is ethnically diverse thus enabling an assessment of the robustness of AutoMitoC across genetic ancestries (S1. Figure 6). Correlations between array and qPCR mtDNA-CN estimates were comparable for individuals of European (N=2431), Latin American (N=1704), African (N=542), South East Asian (N=471), South Asian (N=186), and other ethnic groups (N=360; S1. Figure 6). Bland Altman plots also illustrate the extent of agreement between methods (S1. Figure 7). For every ethnicity, 95% limits of agreement intervals were smaller than expected by chance. Lastly, while all analyses hitherto followed the Mitopipeline condition of requiring > 40,000 autosomal variants for normalization, we observed comparable performance using even 1,000 random rare autosomal probes ($r2=0.60$; $P<5x10^{-300}$) for signal normalization which reduced the runtime from several hours to less than 10 minutes for these 5,791 samples.

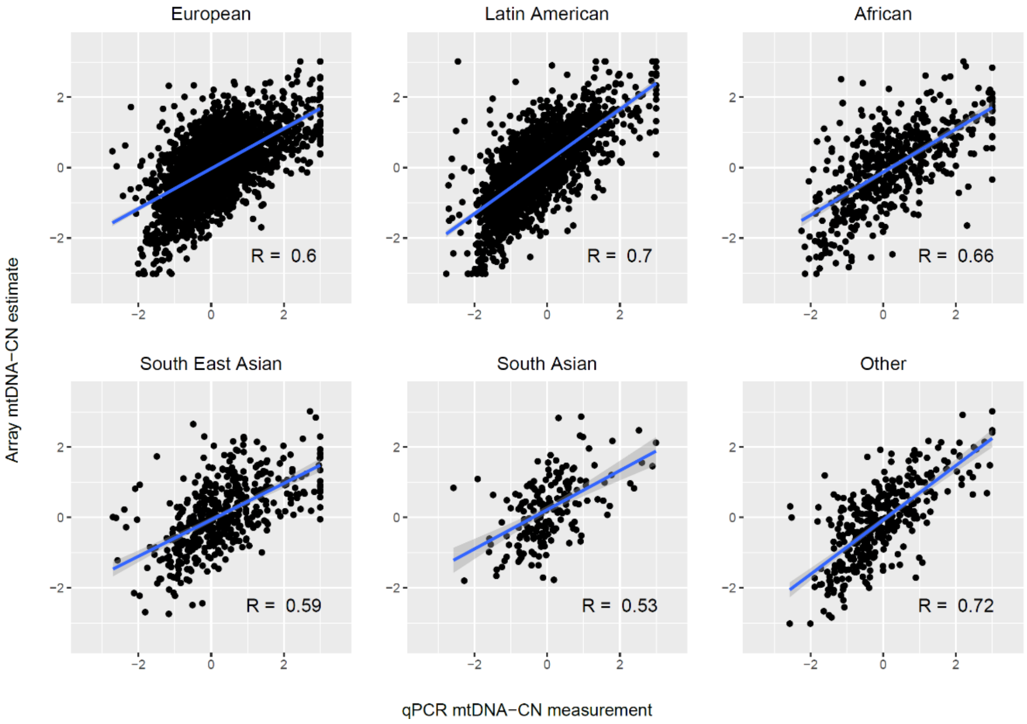


**S1. Figure 6.** Validation of AutoMitoC in an ethnically diverse cohort with qPCR-based estimates. Both qPCR and array-based mtDNA-CN estimates are presented as standardized units (mean=0; SD=1). The sample consisted of 2431 Europeans, 1704 Latin Americans, 542 Africans, 471 South East Asians, 186 South Asians, and 360 participants of other ancestry. Correlations between array and qPCR estimates were comparable for European (r=0.60; $P=2.7x10^{-238}$), Latin American (r=0.70; $P=3.9x10^{-251}$), African (R=0.66; $P=1.8x10^{-68}$), South East Asian (r=0.59; $P=6.2x10^{-46}$), South Asian (r=0.53; $P=4.2x10^{-15}$), and other (r=0.72; $P=5.4x10^{-59}$) ethnic groups. The blue line indicates the linear trendline and the surrounding shaded region indicates the 95% confidence interval for the trendline.

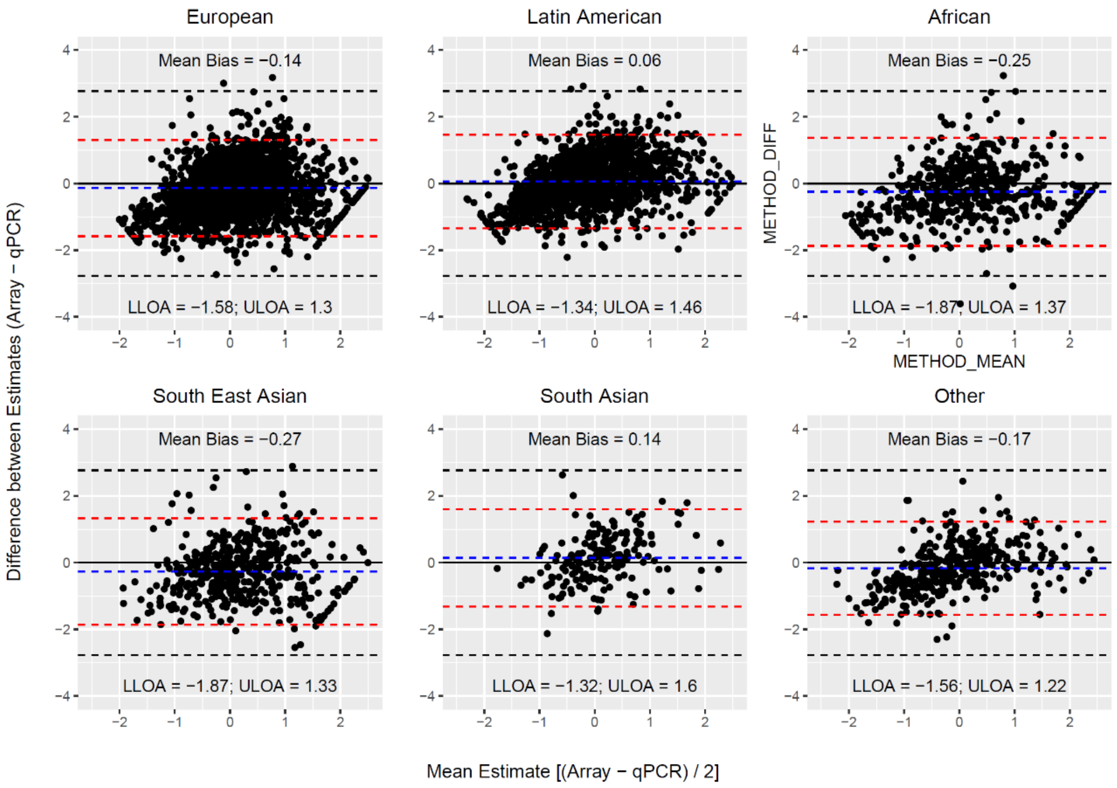


**S1. Figure 7.** Bland Altman plots illustrating the extent of agreement between array and qPCR measurements. The black solid line indicates perfect agreement. The dashed blue line indicates the mean difference (or bias) between estimates. The horizontal red line corresponds to the 95% upper and lower limits of agreement (U/L LOA) for the observed data. The dashed black lines indicate the 95% U/L LOA that is expect under the null for two unrelated variables.

#### Supplementary Methods

*** For all methods and results pertaining to the AutoMitoC pipeline, please see supplementary results I.

##### The UKBiobank study

The UKBiobank is a prospective cohort study including approximately 500,000 UK residents (ages 40-69 years) recruited from 2006-2010 in whom extensive genetic and phenotypic investigations have been and continue to be done(Sudlow et al. 2015). All UKBiobank data reported in this manuscript were accessed through the UKBiobank data showcase under application # 15525. All following analyses described in this supplementary material involve the use of genetic and/or phenotypic data from consenting UKBiobank participants.

#### Genetic Analysis of Common Variants

Data acquisition and quality control

UKBiobank samples were genotyped on either the UK Biobank Array (~450,000) or the UK BiLEVE array (~50,000) for approximately 800,000 variants (Bycroft et al. 2018). Further imputation was conducted by the UKBiobank study team using a combined reference panel of the UK10K and Haplotype Reference Consortium datasets. Imputed genotypes (version 3) for 488,264 UKBiobank participants were downloaded through the European Genome Archive (Category 100319). Samples were removed if they were flagged for any of the UKBiobank-provided quality control annotations (Resource 531; “ukb_sqc_v2.txt”) for high ancestry-specific heterozygosity, high missingness, mismatching genetic ancestry, or sex chromosome aneuploidy (“het.missing.outliers”,“in.white.British.ancestry.subset”, “putative.sex.chromosome.aneuploidy”). Samples were also removed if their submitted gender did not match their genetic sex or if they had withdrawn consent at the time of analysis. Variant quality control consisted of removing variants that had low imputation quality (INFO score $\leq$ 0.30), were rare (MAF $\leq$ 0.005), or were in Hardy Weinberg Disequilibrium (HWE P $\leq 1x10^{-10}$). The HWE test was conducted within a subset of unrelated individuals for each ethnic strata, though all related individuals were retained for subsequent GWAS analysis. Lastly, in special consideration of mtDNA-CN as the GWAS phenotype, we also removed variants within “NUMTs”, which refer to regions of the nuclear genome that exhibit homology to the mitochondrial genome due to past transposition of mitochondrial sequences. Accordingly, NUMTs represent a specific confounder of mtDNA-CN GWAS analyses which may lead to false positive associations. NUMT boundaries were obtained from the UCSC NumtS Sequence (numtSeq) track, which is based on the Reference Human NumtS curated by Simone *et al.* (2011) (Simone et al. 2011). All sample and variant quality control of imputed genotypes were executed using qctools and resultant bgen files were indexed with bgenix. Smaller ethnic groups with similar genetic ancestry were consolidated; individuals self-reporting as “African” or “Caribbean” were combined into a larger “African” stratum and individuals self-reporting as “Indian” or “Pakistani” were combined into a “South Asian” stratum. After quality control, 359689 British, 10598 Irish, 13189 Other White, 6172 South Asian, and 6133 African samples passing quality control also had suitable array-based mtDNA-CN estimates for subsequent GWAS analyses.

Genome-wide association study (GWAS)

GWAS were initially conducted in an ethnicity-stratified manner. The number of variants tested for association with mtDNA-CN varied for British (M=10,728,525), Irish (M=10,707,537), Other White (M=10,894,497), South Asian (M=11,350,981), and African (M=18,981,896) study participants, respectively. GWAS was performed using the REGENIE framework which consists of two steps (Mbatchou et al. 2020). In step 1, mtDNA-CN was predicted using a ridge regression model fit on a set of high-quality genotyped SNPs (MAF>0.01, MAC>100, genotype and sample missingness above 10%, and passing HWE ($p>10^{-15}$)) across the whole genome in blocks of 1000 SNPs. In step 2, the linear regression model was used to test the association of all SNPs adjusting for age, $age^2$, sex, chip type, 20 genetic principal components, and blood cell counts (white blood cell, platelet, and neutrophil counts), and conditional on the model from step 1.

Blood cell counts were determined for blood specimen collected at the initial assessment visit using Beckman Coulter LH750 analyzers (https://biobank.ndph.ox.ac.uk/showcase/showcase/docs/haematology.pdf). Information on blood cell counts was retrieved from the UKBiobank data showcase. Individuals with missing values for any blood cell counts (~2.5%) were removed from any subsequent analysis involving blood cell counts. Quality control of blood counts was done following the same procedure as Longchamps *et al.* (2019) (Longchamps 2019). Except for platelet counts, all blood cell counts were log-transformed and samples exhibiting outlying values were removed (~4% samples). Lastly, values were standardized to have a mean of 0 and standard deviation of 1.

After ethnicity-specific GWAS were performed, results were combined through meta-analysis using METAL (Willer, Li, and Abecasis 2010). European (N=383,476) and trans-ethnic (N=395,718) GWAS meta-analyses were performed. We found that results from the trans-ethnic meta-analyses strongly resembled that of the European meta-analysis due to the high proportion of Europeans (97%). Accordingly, we report results from the European meta-analysis as the primary GWAS. To summarize statistical associations, Manhattan plots and quantile-quantile plots were generated by uploading summary statistics into the locus zoom web platform (https://my.locuszoom.org/) (Pruim et al. 2010). LD-score regression was performed to calculate the LD-score intercept by uploading GWAS results to the LDhub test center (http://ldsc.broadinstitute.org/) (Bulik-Sullivan et al. 2015). As per the instructions, all variants within the MHC region on chromosome 6 were removed prior to uploading. Annovar (version date 2020-06-07) was used to functionally annotate genome-wide significant loci based on their proximity (+/- 250kb) to genes (RefSeq), predicted effect on amino acid sequence, allele frequency in external datasets (1000Genomes), clinical pathogenicity (Clinvar), and in silico deleteriousness (CADD), and eQTL information (GTEx v8) (Abecasis et al. 2012; GTEX 2014; Landrum et al. 2015; Rentzsch et al. 2019).

*Fine-mapping of GWAS signals*

We followed a similar protocol to Vuckovic *et al.* (2020) for fine-mapping mtDNA-CN loci (Vuckovic et al. 2020). All 9,602 genome-wide significant variants were consolidated into genomic blocks by grouping variants within 250kb of each other, yielding 72 distinct genomic blocks. LDstore was used to compute a pairwise LD correlation matrix for all variants within each block and across all samples included in the European GWAS meta-analysis (Benner et al. 2017). For each genomic block, FINEMAP was used to perform stepwise conditional regression, leading to 88 conditionally independent variants at genome-wide significance (Benner et al. 2016). The number of conditionally independent genetic signals per genomic block was used to inform the subsequent fine-mapping search parameters. Finally, the FINEMAP random stochastic search algorithm was applied to derive 95% credible sets constituting candidate causal variants that jointly contributed to 95% (or higher) of the posterior inclusion probabilities (Benner et al. 2016).

*Mitochondrial expression quantitative trait loci (mt-eQTL)*

Among our GWAS hits, we searched for mt-eQTLs using information from Ali *et al*. (2019), “Nuclear genetic regulation of the human mitochondrial transcriptome” (Ali et al. 2019). All variants in both Tables 1 and 2 were queried in the mtDNA-CN summary statistics. When mt-eQTLs also had reported effect estimates, the consistency in direction-of-effects between mt-eQTL and mtDNA-CN associations was reported (S2. Table 3).

*Gene prioritization & pathway analyses*

The Data-driven Expression-prioritized Integration for Complex Traits (DEPICT) v.1.1 tool was used to map mtDNA-CN loci to genes based on shared co-regulation of gene expression (Pers et al. 2015). Genome-wide significant variants from the European GWAS meta-analysis were "clumped" into independent loci using PLINK "--clump-p1 5e-8 --clump-kb 500 --clump-r2 0.05" with LD correlation matrix derived from 1000Genomes Europeans (Purcell et al. 2007). DEPICT was subsequently run on independent SNPs using default settings. DEPICT identified 91 genes in total at a FDR of 0.05. Of the 91 genes, 4 non-coding genes were excluded from subsequent analyses for lack of a match in the GeneMANIA database(Warde-Farley et al. 2010). The excluded genes include a pseudogene (*PTMAP3*), an intronic transcript (*ALMS1-IT1*), and 2 long non-coding RNAs (*SNHG15*, *RP11-125K10.4*). The remaining 87 DEPICT-prioritized genes were uploaded to the GeneMANIA web platform (https://genemania.org/), which mines publicly available biological datasets to identify additional related genes based on functional associations (genetic interactions, pathways, co-expression, co-localization and protein domain homology). Based on the combined list of DEPICT and GeneMANIA identified genes, a network was formed in GeneMANIA maximizing the connectivity between all input genes using the default "Assigned based on query gene" setting to weight the network. Functional enrichment analysis was then performed to identify overrepresented Gene Ontology (GO) terms among all network genes (Gene and Consortium 2000). All network genes with at least one GO annotation were compared to a background comprising all GeneMANIA genes with GO annotations.

*Mitochondrial annotation-based analyses*

To complement the previous analyses, we labelled prioritized genes with MitoCarta3 annotations and performed subsequent statistical enrichment analyses (Rath et al. 2021). MitoCarta3 is an exquisite database of mitochondrial protein annotations, which draws from mass spectrophotometry and GFP colocalization experiments of isolated mitochondria from 14 different tissues, as well as a plethora of other sources including literature review, to assign all human genes statuses indicating whether the corresponding proteins are expressed in the mitochondria or not. We tested whether prioritized genes were enriched for the mitochondrial proteome by using a binomial test in R. The number of "trials" was set to the total number of DEPICT and GeneMANIA-prioritized genes (107); the number of "successes" was set to the aforementioned gene subset that were labelled as mitochondrial proteins by MitoCarta3 (27); finally, the expected probability was set to the number of nuclear-encoded MitoCarta3 genes divided by the total number of genes (1120/18922). Furthermore, a t-test was used to compare mean PGC-1A induced fold change for the 27 genes as compared to the mean PGC-1A induced fold change for all 1120 nuclear MitoCarta3-annotated genes. MitoCarta3 genes with missing values were excluded from this analysis. Lastly, the 27 genes were labelled based on MitoCarta3 "MitoPathways". Only the top level pathway (i.e. parent node) was ascribed to each gene within the main text though detailed pathway annotations are available S2. Table 5.

**Genetic Analysis of Rare Variants**

*Data acquisition and quality control*

Population-level whole-exome sequencing (WES) variant genotypes (UKB data field: 23155) for 200,643 UKBiobank participants corresponding to 17,975,236 variants were downloaded using the gfetch utility. These data represent the second tranche of WES data released by the UKBiobank and differs from the first tranche (~50K samples) which was used for the development of the AutoMitoC pipeline. Quality control of WES data was conducted as follows. First, 11 samples who withdrew consent by the time of analysis were removed. Second, 83,700 monomorphic variants were removed. Third, 369,215 variants with non-missing genotypes present in less than 90% of samples were removed. Fourth, 2 samples with call rates less than 99% were removed. Fifth, 18 samples exhibiting discordance between genetic and reported sex were removed. Sixth, through visual inspection of scatterplots of the first two genetic principal components, 3 outlying samples whose locations strongly departed from their putative ethnicity cluster were removed. Seventh, 35,317 variants deviating from Hardy Weinberg Equilibrium were removed. Eighth, 12,765 samples belonging to smaller ethnic groups with less than 5000 samples (South Asian=3395; African=3168; Other=6,202) were removed. Ninth, we selected for a maximal number of unrelated samples and excluded 14,156 samples exhibiting third degree or closer relatedness. Finally, 12,394,404 non-coding variants were removed, and 5,176,300 protein-altering variants (stopgain, stoploss, startloss, splicing, missense, frameshift and in-frame indels) were retained in 173,688 samples.

*Exome-wide association testing to identify rare mtDNA-CN loci*

Of the 173,688 individuals passing quality control, 147,740 had non-missing mtDNA-CN estimates. Further variant inclusion criteria were implemented: variants that were rare (MAF $\leq$ 0.001), non-synonymous, and predicted to be clinically deleterious by Mendelian Clinically Applicable Pathogenicity (M-CAP) v.1.4 scores (or were highly disruptive variant types including frameshift indel, stopgain, stoploss, or splicing) were retained (Jagadeesh et al. 2016). Herein, such variants are referred to as "rare variants" for simplicity. For each gene, rare allele counts were added per sample. 18,890 genes with a total minor allele count of least 10 were subsequently analyzed (exome-wide significance $P < 0.05/18890 = 2.65 \times 10^{-6}$). Linear regression was conducted using mtDNA-CN as the dependent variable and the rare alleles counts per gene as the independent variable. The same set of covariates used in the primary GWAS were also employed in this analysis.

*Phenome-wide association testing for rare SAMHD1 mutation carrier status*

To identify disease phenotypes associated with carrying a rare *SAMHD1* mutation, we maximized sample size for phenome-wide association testing by analyzing the larger set of 173,688 WES samples (with or without suitable mtDNA-CN estimates). Disease outcomes were defined using the previously published "PheCode" classification scheme to aggregate ICD-10 codes from hospital episodes (field ID 41270), death registry (field ID 40001 and 40002), and cancer registry (field ID 40006) records(Denny et al. 2013; Wu et al. 2019). Further manual review was performed to exclude cases of sex-specific outcomes that may be erroneously attributed to the opposite genetic sex. Logistic regression was applied to test the association of *SAMHD1* mutation carrier status versus 771 PheCodes (phenome-wide significance $P < 0.05/771 = 6.49 \times 10^{-5}$) with a minimal case sample size of 300 (Wei et al. 2017). The same set of covariates used in the primary GWAS were also employed in this analysis.

**Mendelian Randomization Analysis**

*Disease Outcomes*

To assess evidence for a causal role of mtDNA-CN on mitochondrial disorder-related traits, we first defined a list of testable disease outcomes related to mitochondrial disorders. 36 clinical manifestations from a review paper by Gorman *et al.* (2016) were cross-referenced to GWAS traits analyzed by the FinnGen consortium (Gorman et al. 2016; Madewell et al. 2020). The

FinnGen consortium is a collaborative research entity aggregating genomic data from 9 Finnish biobanks with phenotypic data from electronic health records (https://finngen.gitbook.io/documentation/data-description#summary-association-statistics). FinnGen GWAS (v4) have been performed for 176,899 participants and 2,444 disease endpoints using the SAIGE method which entails a logistic mixed model with saddle point approximation to account for imbalanced case-control ratios(Zhou et al. 2018). Of these 2,444 disease endpoints, 10 traits corresponded to one of the 36 clinical manifestations of mitochondrial disease and had a case prevalence greater than 1% in FinnGen including type 2 diabetes (N=23,364), mood disorder (N=20,288), sensorineural hearing loss (N=12,550), cerebrovascular disease (N=10,367), migraine (N=6,687), dementia (5,675), epilepsy (N=4,558), paralytic ileus and intestinal obstruction (N=2,999), and cardiomyopathy (N=2,342). Genome-wide summary statistics were downloaded for these 10 traits, from which effect estimates and standard errors were used in subsequent Mendelian randomization analyses to define the effect of selected genetic instruments on disease risk.

*Genetic Instrument Selection*

First, genome-wide significant variants from the European GWAS meta-analysis of mtDNA-CN were chosen (N=383476). Second, we matched these variants to the FinnGen v4 GWAS datasets (Madewell et al. 2020). Third, to enrich for variants that directly act through mitochondrial processes, we only retained those within 100kb of genes encoding for proteins that localize to the mitochondria based on MitoCarta3 annotations (Rath et al. 2021). Fourth, we performed LD-pruning in PLINK with 1000Genomes Europeans as the reference panel to ascertain an independent set of genetic variants (LD r2 > 0.01), resulting in 34 variants (Abecasis et al. 2012; Purcell et al. 2007). Lastly, to mitigate potential for horizontal pleiotropy, we further removed variants with strong evidence of acting through alternative pathways by performing a phenome-wide search across published GWAS with Phenoscanner V2 (Kamat et al. 2019). Variants strongly associated with other phenotypes ($P<5x10^{-20}$) were removed unless the variant was a coding mutation located within gene encoding for the mitochondrial proteome (MitoCarta3) or had an established mitochondrial role based on manual literature review (Rath et al. 2021). Seven genetic variants were removed based on these criteria including rs8067252 (*ADAP2*), rs56069439 (*ANKLE1*), rs2844509 (*ATP6V1G2-DDX39B*), rs73004962 (*PBX4*), rs7412 (*APOE*), rs385893 (*AK3, RCL1*), and rs1613662 (*GP6*) (S2. Table 8).

*Mendelian Randomization & Sensitivity Analyses*

Two sample Mendelian Randomization analyses were performed using the “TwoSampleMR” and “MRPRESSO” R packages (Hemani et al. 2018; Verbanck et al. 2018).  Effect estimates and standard errors corresponding to the 27 genetic variants on mtDNA-CN (exposure) and mitochondrial disease phenotypes (outcome) were derived from the European GWAS meta-analysis and FinnGen v4 GWAS summary statistics, respectively (S2. Table 9). Three MR methodologies were employed including Inverse Variance Weighted (primary method), Weighted Median, and MR-EGGER methods. MR-PRESSO was used to detect global heterogeneity and P-values were derived based on 1000 simulations. If significant global heterogeneity was detected ($P<0.05$), a local outlier test was conducted to detect outlying SNPs. After removal of outlying SNPs, MR analyses were repeated. In the absence of heterogeneity (Egger-intercept $P \geq 0.05$; MR-PRESSO global heterogeneity $P \geq 0.05$), we reported the inverse-variance weighted result. In the presence of balanced pleiotropy (MR-PRESSO global heterogeneity $P < 0.05$) and absence of directional pleiotropy (Egger-intercept $P \geq 0.05$), we reported the weighted median result. In the presence of directional pleiotropy (Egger-intercept $P < 0.05$), we reported the MR-EGGER result. We also performed the Steiger directionality test to ensure that a greater proportion of variance in mtDNA-CN was explained than risk of the outcome.

#### Supplementary Results II

##### Supplementary Table Titles

**S2. Table 1**. Annotated genome-wide significant mtDNA-CN loci

**S2. Table 2**. FINEMAP results for 72 mtDNA-CN loci and 82 independent signals

**S2. Table 3**. Overlap between mtDNA-CN loci and MT-eQTLs (Ali *et al.*, 2019)

**S2. Table 4**. DEPICT gene prioritization results

**S2. Table 5**. MitoCarta3 annotations for DEPICT and GeneMANIA-prioritized genes

**S2. Table 6.** Rare variant exome-wide association testing for mtDNA-CN loci

**S2. Table 7.** Rare variant SAMHD1 phenome-wide association testing with disease status

**S2. Table 8**. Phenoscanner search results for genetic variants initially considered as genetic instruments for Mendelian Randomization analysis

**S2. Table 9**. Mendelian Randomization analyses of mtDNA-CN versus mitochondrial disease phenotypes

#### Supplementary Figures

**African** **South Asian** **Other White**

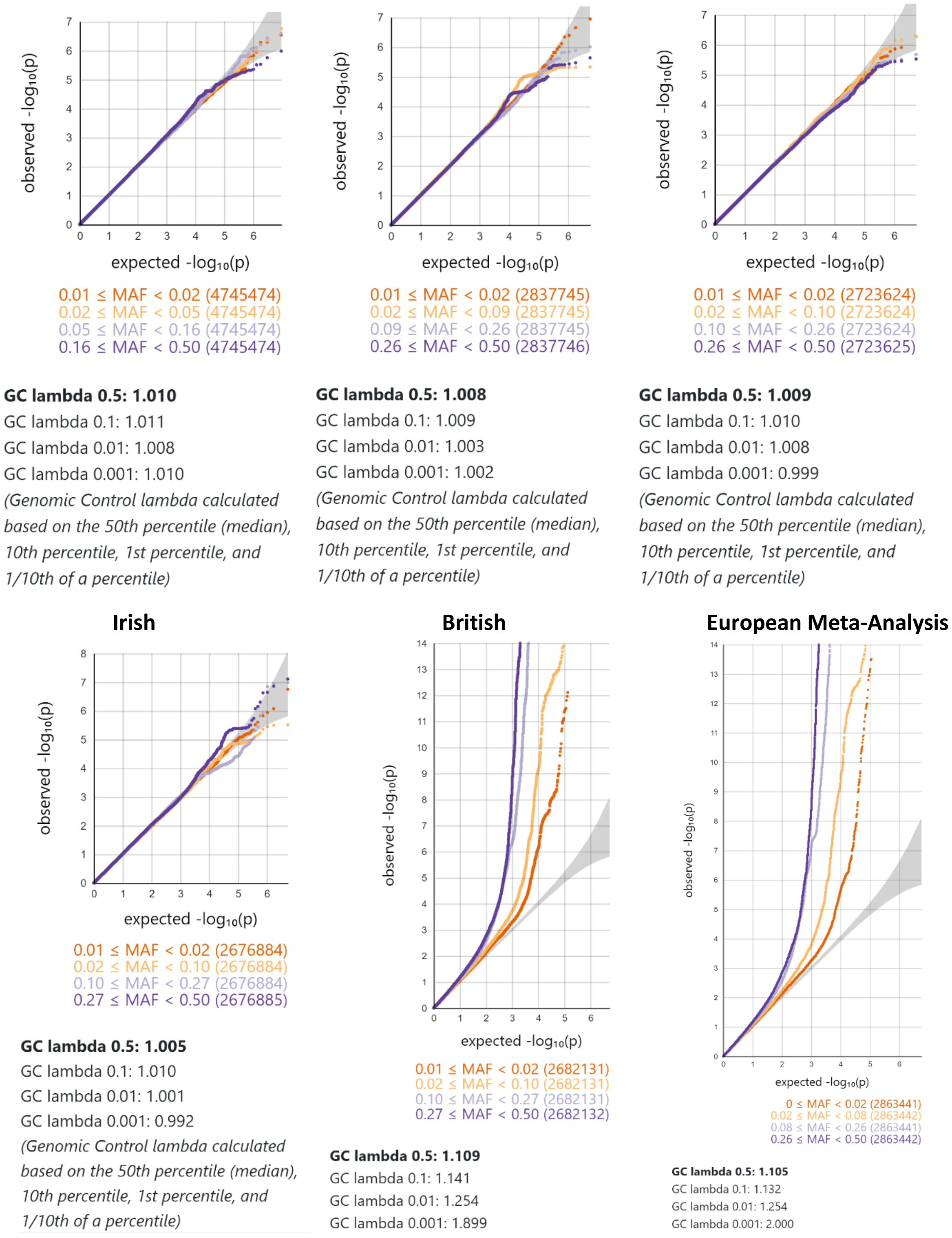


**S2. Figure 1**. MAF and ethnicity-stratified GWAS quantile-quantile plots.

**S2. Figure 2 (Extended Figures)**. Locus zoom plots for 72 loci and 82 conditionally independent genetic signals. Variants with the highest fine-mapping posterior probability are labelled by their genomic coordinates (GRCh37). Pairwise correlation between the lead variant and proximal variants were colour-coded based on the 1000Genomes Europeans reference panel.

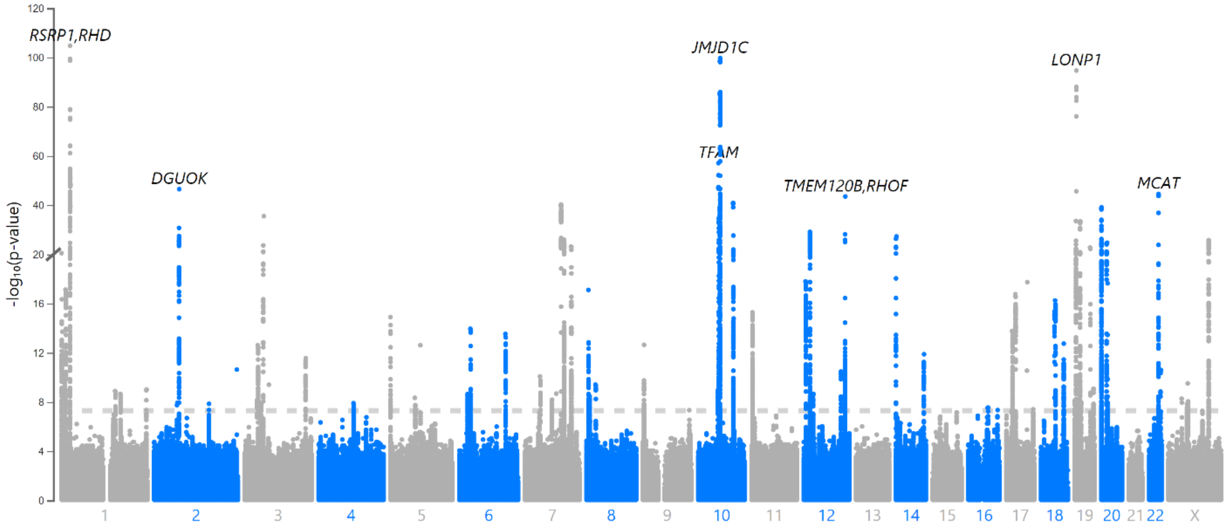


**S2 Figure 3.** Manhattan plot for trans-ethnic GWAS meta-analysis (N=395,781).

A B

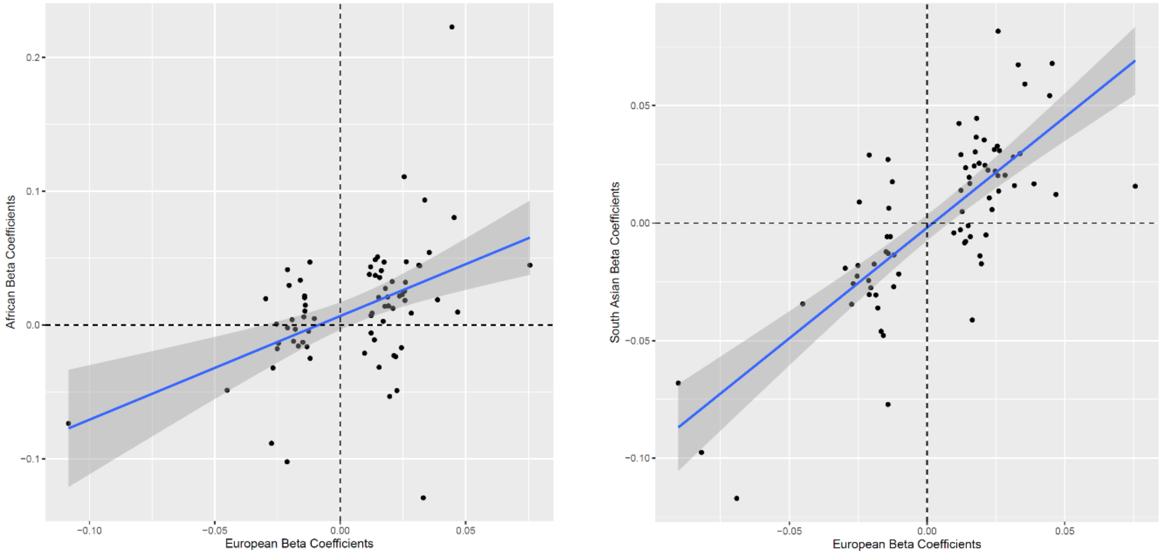


**S2 Figure 4.** Correlation between conditionally independent mtDNA-CN loci effect estimates derived from European GWAS meta-analyses (x-axes) vs. effect estimates from Non-European GWAS (y-axes). Comparisons for African (A) and South Asian (B) GWAS analyses are presented. Of the total 82 conditionally independent signals identified using the European GWAS meta-analysis, 73 and 75 variants were available for comparison in African and South Asian GWAS, respectively.

A B

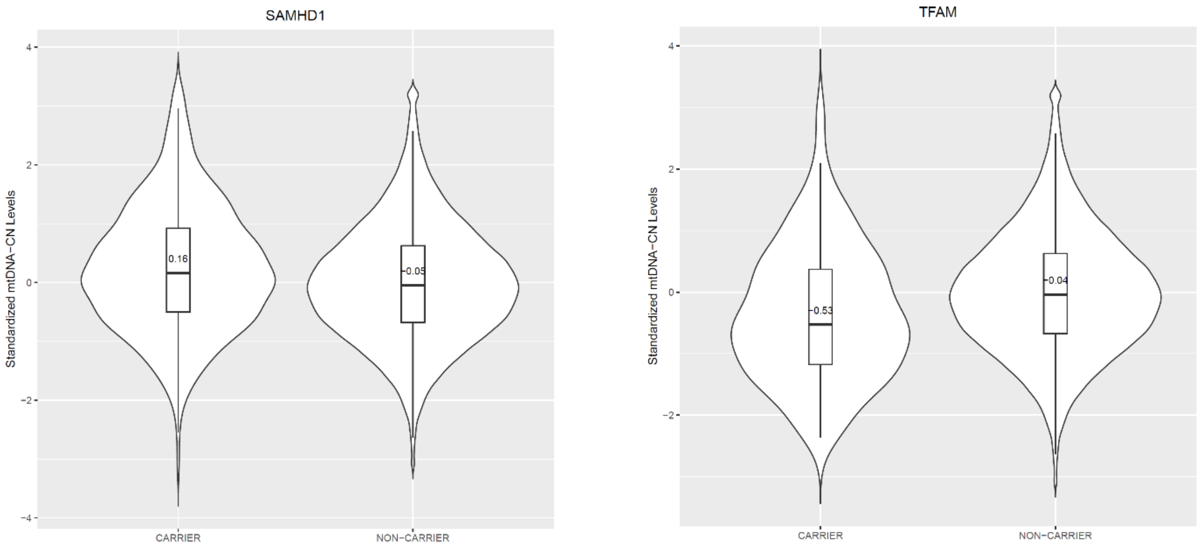


**S2. Figure 5**. Violin plots showing the distribution of mtDNA-CN for carriers and non-carriers of (A) *SAMHD1* and (B) *TFAM* rare nonsynonymous and deleterious (MCAP > 0.025) variants.

#### References

Abecasis, Goncalo R et al. 2012. “An Integrated Map of Genetic Variation from 1,092 Human Genomes.” *Nature* 491(7422): 56–65. http://www.pubmedcentral.nih.gov/articlerender.fcgi?artid=3498066&tool=pmcentrez&rendertype=abstract (July 9, 2014).

Ali, Aminah T. et al. 2019. “Nuclear Genetic Regulation of the Human Mitochondrial Transcriptome.” *eLife* 8: 1–23.

Altschul, S F et al. 1990. “Basic Local Alignment Search Tool.” *Journal of molecular biology* 215(3): 403–10. http://www.ncbi.nlm.nih.gov/pubmed/2231712.

Ashar, Foram N et al. 2017. “Association of Mitochondrial DNA Copy Number With Cardiovascular Disease.” 21205(11): 1247–55.

Benner, Christian et al. 2016. “FINEMAP: Efficient Variable Selection Using Summary Data from Genome-Wide Association Studies.” *Bioinformatics* 32(10): 1493–1501.

———. 2017. “Prospects of Fine-Mapping Trait-Associated Genomic Regions by Using Summary Statistics from Genome-Wide Association Studies.” *The American Journal of Human Genetics* 101(4): 539–51. https://linkinghub.elsevier.com/retrieve/pii/S0002929717303348.

Bulik-Sullivan, Brendan et al. 2015. “LD Score Regression Distinguishes Confounding from Polygenicity in Genome-Wide Association Studies.” *Nature Genetics* 47(3): 291–95.

Bycroft, Clare et al. 2018. “The UK Biobank Resource with Deep Phenotyping and Genomic Data.” *Nature* 562(7726): 203–9. http://www.nature.com/articles/s41586-018-0579-z.

Denny, Joshua C et al. 2013. “Systematic Comparison of Phenome-Wide Association Study of Electronic Medical Record Data and Genome-Wide Association Study Data.” *Nature biotechnology* 31(12): 1102–10. http://www.ncbi.nlm.nih.gov/pubmed/24270849.

Diskin, Sharon J et al. 2008. “Adjustment of Genomic Waves in Signal Intensities from Whole-Genome SNP Genotyping Platforms.” *Nucleic acids research* 36(19): e126. http://www.ncbi.nlm.nih.gov/pubmed/18784189.

Fazzini, Federica et al. 2019. “Mitochondrial DNA Copy Number Is Associated with Mortality and Infections in a Large Cohort of Patients with Chronic Kidney Disease.” 8: 480–88.

Gene, The, and Ontology Consortium. 2000. “Gene Ontology : Tool for The.” 25(may): 25–29.

Gorman, Gráinne S. et al. 2016. “Mitochondrial Diseases.” *Nature Reviews Disease Primers* 2.

GTEX. 2014. “The Genotype-Tissue Expression (GTEx) Project The.” *Nature Genetics*

45(6): 580–85.

Harris, Charles R. et al. 2020. “Array Programming with NumPy.” *Nature* 585(7825): 357–62. http://www.nature.com/articles/s41586-020-2649-2.

Hemani, Gibran et al. 2018. “The MR-Base Platform Supports Systematic Causal Inference across the Human Phenome.” *eLife* 7: 1–29.

Jagadeesh, Karthik A et al. 2016. “M-CAP Eliminates a Majority of Variants of Uncertain Significance in Clinical Exomes at High Sensitivity.” *Nature genetics* 48(12): 1581–86. http://www.ncbi.nlm.nih.gov/pubmed/27776117.

Kamat, Mihir A et al. 2019. “PhenoScanner V2: An Expanded Tool for Searching Human Genotype-Phenotype Associations.” *Bioinformatics (Oxford, England)* 35(22): 4851–53. http://www.ncbi.nlm.nih.gov/pubmed/31233103.

Landrum, M. J. et al. 2015. “ClinVar: Public Archive of Interpretations of Clinically Relevant Variants.” *Nucleic Acids Research*. http://nar.oxfordjournals.org/lookup/doi/10.1093/nar/gkv1222.

Lane, John. 2014. “MitoPipeline: Generating Mitochondrial Copy Number Estimates from SNP Array Data in Genvisis.” http://genvisis.org/MitoPipeline/.

Longchamps, Ryan Joseph. 2019. “EXPLORING THE ROLE OF MITOCHONDRIAL DNA QUANTITY AND QUALITY.” *Biorxiv* (August).

Madewell, Zachary J et al. 2020. “Findings and Insights from the Genetic Investigation of Age of First Reported Occurrence for Complex Disorders in the UK Biobank and FinnGen.” *medRxiv*: 1–13.

Mbatchou, Joelle et al. 2020. “Computationally Efficient Whole Genome Regression for Quantitative and Binary Traits.” : 1–88.

O’Donnell, Martin J et al. 2010. “Risk Factors for Ischaemic and Intracerebral Haemorrhagic Stroke in 22 Countries (the INTERSTROKE Study): A Case-Control Study.” *Lancet* 376(9735): 112–23. http://www.ncbi.nlm.nih.gov/pubmed/20561675 (July 11, 2014).

Pers, Tune H. et al. 2015. “Biological Interpretation of Genome-Wide Association Studies Using Predicted Gene Functions.” *Nature Communications* 6: 5890. http://www.pubmedcentral.nih.gov/articlerender.fcgi?artid=4420238&tool=pmcentrez&rendertype=abstract.

Pruim, Randall J et al. 2010. “LocusZoom : Regional Visualization of Genome-Wide Association Scan Results.” 26(18): 2336–37.

Purcell, Shaun et al. 2007. “PLINK: A Tool Set for Whole-Genome Association and Population-Based Linkage Analyses.” *American journal of human genetics* 81(3): 559–75.

Rath, Sneha et al. 2021. “MitoCarta3.0: An Updated Mitochondrial Proteome Now with Sub-Organelle Localization and Pathway Annotations.” *Nucleic acids research*

49(D1): D1541–47.

Rentzsch, Philipp et al. 2019. “CADD: Predicting the Deleteriousness of Variants throughout the Human Genome.” *Nucleic Acids Research* 47(D1): D886–94. https://academic.oup.com/nar/article/47/D1/D886/5146191.

Simone, Domenico et al. 2011. “The Reference Human Nuclear Mitochondrial Sequences Compilation Validated and Implemented on the UCSC Genome Browser.” *BMC genomics* 12: 517. http://www.ncbi.nlm.nih.gov/pubmed/22013967.

Sudlow, Cathie et al. 2015. “UK Biobank: An Open Access Resource for Identifying the Causes of a Wide Range of Complex Diseases of Middle and Old Age.” *PLoS medicine* 12(3): e1001779. http://www.ncbi.nlm.nih.gov/pubmed/25826379.

Verbanck, Marie, Chia Yen Chen, Benjamin Neale, and Ron Do. 2018. “Detection of Widespread Horizontal Pleiotropy in Causal Relationships Inferred from Mendelian Randomization between Complex Traits and Diseases.” *Nature Genetics* 50(5): 693–98. http://dx.doi.org/10.1038/s41588-018-0099-7.

Virtanen, Pauli et al. 2020. “SciPy 1.0: Fundamental Algorithms for Scientific Computing in Python.” *Nature Methods* 17(3): 261–72. http://www.nature.com/articles/s41592-019-0686-2.

Vuckovic, Dragana et al. 2020. “The Polygenic and Monogenic Basis of Blood Traits and Diseases.” *Cell* 182(5): 1214-1231.e11. http://www.ncbi.nlm.nih.gov/pubmed/32888494.

Wang, Kai et al. 2007. “PennCNV: An Integrated Hidden Markov Model Designed for High-Resolution Copy Number Variation Detection in Whole-Genome SNP Genotyping Data.” *Genome research* 17(11): 1665–74. http://www.ncbi.nlm.nih.gov/pubmed/17921354.

Warde-Farley, David et al. 2010. “The GeneMANIA Prediction Server: Biological Network Integration for Gene Prioritization and Predicting Gene Function.” *Nucleic Acids Research* 38(SUPPL. 2): 214–20.

Wei, Wei Qi et al. 2017. “Evaluating Phecodes, Clinical Classification Software, and ICD-9-CM Codes for Phenome-Wide Association Studies in the Electronic Health Record.” *PLoS ONE* 12(7): 1–16.

Willer, Cristen J., Yun Li, and Gonçalo R. Abecasis. 2010. “METAL: Fast and Efficient Meta-Analysis of Genomewide Association Scans.” *Bioinformatics* 26(17): 2190–91.

Wu, Patrick et al. 2019. “Mapping ICD-10 and ICD-10-CM Codes to Phecodes: Workflow Development and Initial Evaluation.” *JMIR medical informatics* 7(4): e14325. http://www.ncbi.nlm.nih.gov/pubmed/31553307.

Zhang, Yiyi et al. 2017. “Association between Mitochondrial DNA Copy Number and Sudden Cardiac Death : Findings from the Atherosclerosis Risk in Communities Study ( ARIC ).” : 3443–48.

Zhou, Wei et al. 2018. “Efficiently Controlling for Case-Control Imbalance and Sample Relatedness in Large-Scale Genetic Association Studies.” *Nature Genetics* 50(9): 1335–41. http://www.nature.com/articles/s41588-018-0184-y.
